# Genetic evidence supports the development of SLC26A9 targeting therapies for the treatment of lung disease

**DOI:** 10.1101/2021.10.07.21264392

**Authors:** Jiafen Gong, Gengming He, Cheng Wang, Claire Bartlett, Naim Panjwani, Scott Mastromatteo, Fan Lin, Katherine Keenan, Julie Avolio, Anat Halevy, Michelle Shaw, Mohsen Esmaeili, Guillaume Côté-Maurais, Damien Adam, Stéphanie Bégin, Candice Bjornson, Mark Chilvers, Joe Reisman, April Price, Michael Parkins, Richard Van Wylick, Yves Berthiaume, Lara Bilodeau, Dimas Mateos-Corral, Daniel Hughes, Mary J. Smith, Nancy Morrison, Janna Brusky, Elizabeth Tullis, Anne L. Stephenson, Bradley S. Quon, Pearce Wilcox, Winnie M. Leung, Melinda Solomon, Lei Sun, Emmanuelle Brochiero, Theo J. Moraes, Tanja Gonska, Felix Ratjen, Johanna M. Rommens, Lisa J. Strug

## Abstract

**Background:** Over 400 variants in the cystic fibrosis (CF) transmembrane conductance regulator (*CFTR*) are CF-causing. CFTR modulators target variants to improve lung function, but marked variability in response exists and current therapies do not address all CF-causing variants highlighting unmet needs. Alternative epithelial ion channel/transporters such as SLC26A9 could compensate for CFTR dysfunction, providing therapeutic targets that may benefit all individuals with CF.

**Method:** We investigate the relationship between rs7512462, a marker of *SLC26A9* activity, and lung function pre- and post-treatment with CFTR modulators in Canadian and US CF cohorts, in the general population, and in those with chronic obstructive pulmonary disease (COPD).

**Results:** Rs7512462 CC genotype is associated with greater lung function in CF individuals with minimal function variants (for which there are currently no approved therapies; p=0.008); and for gating (p=0.033) and p.Phe508del/ p.Phe508del (p=0.006) genotypes upon treatment with CFTR modulators. In parallel, human nasal epithelia with CC and p.Phe508del/p.Phe508del after Ussing chamber analysis of a combination of approved and experimental modulator treatments show greater CFTR function (p=0.0022). Beyond CF, rs7512462 is associated with lung function in a meta-analysis of the UK Biobank and Spirometa Consortium (min p=2.74×0^-44^) and provides p=0.0891 in an analysis of COPD case-control status in the UK Biobank defined by spirometry.

**Conclusion:** These findings support SLC26A9 as a therapeutic target to improve lung function for all people with CF and in individuals with other obstructive lung diseases.

## Background

Cystic Fibrosis [CF (MIM:219700)] is a common life-limiting autosomal recessive genetic disease caused by pathogenic variants in the cystic fibrosis transmembrane conductance regulator (CFTR; MIM:602421). CF affects multiple organs; morbidity in the pancreas and intestine are seen at birth (1-3), while progressive lung disease is experienced throughout the course of disease and is the major cause of morbidity and mortality. Variability in disease severity across the affected organs is due, in part, to variation in other genes, referred to as modifier genes. Modifier genes impact phenotypic expressivity in the presence of a dysfunctional major causal gene, for example, *CFTR* (1, 4-7).

CFTR is localized to the cell membrane of epithelial cells and functions as an ion channel. Over 400 variants have been reported to cause CF through variable effects on protein function (http://cftr2.org; (8)). CF-causing mutations impact apical chloride and bicarbonate transport, altering hydration and pH of airway surface liquid resulting in viscous mucus. Accumulation of this viscous mucus leads to cycles of infection and inflammation, obstructing and damaging the airways, resulting in progressive lung damage and end-stage lung disease (9).

Developments in CF therapeutics over the past decade have been transformative, altering the management paradigm from treating the downstream consequences of dysfunctional CFTR and delaying the progression of the disease, to treating the basic defect in an individual’s CFTR itself: *precision medicine*. New drugs enhance CFTR function by directly targeting the different defects in the protein. For example, individuals with gating mutations have access to a highly effective modulator, ivacaftor (IVA), which is a potentiator that increases the opening probability of CFTR to aid chloride and bicarbonate ion transport in CF epithelia (10). The most common CF causing allele Phe508del (c.1521_1523delCTT; p.Phe508del) (11) displays minimal CFTR at the apical membrane due to processing defects (5, 6), and once at the cell surface the Phe508del protein exhibits reduced opening probability and stability. Pharmaceuticals targeting this defect include a combination therapy of IVA and a CFTR corrector lumacaftor (LUM) or tezacaftor (TEZ) that improves the Phe508del processing to increase cell surface localized protein as well as channel gating. More recently, a triple combination therapy of another corrector, elexacaftor, combined with tezacaftor and ivacaftor (ETI) has been approved in the United States (US) for individuals with at least one Phe508del aged 6 and above or other mutations responsive to Trikafta (https://www.cff.org/Trials/Pipeline/details/10150/Elexacaftor-tezacaftor-ivacaftor-Trikafta). Elexacaftor stabilizes the protein within the cell membrane, resulting in greater improvements in lung function over LUM/IVA or TEZ/IVA alone (12, 13). ETI is now indicated by the FDA for 90% of individuals with CF (https://www.fda.gov/news-events/press-announcements/fda-approves-new-breakthrough-therapy-cystic-fibrosis).

Although significant progress has been made in the development of pharmaceuticals for precision medicine in CF, several challenges remain. First, not all individuals with *CFTR* genotypes for which eligible pharmaceuticals are available respond to those treatments. Second, for those who do show a positive improvement in lung function, there is significant variability in the response (12-15), which could be augmented with additional therapies. Third, there are loss of function alleles that cannot be addressed using the current paradigm, and therefore an alternative approach to therapy beyond potentiators and correctors of CFTR is necessary.

Several alternative approaches are being actively pursued, including CFTR gene restoration and/or correction or alternative targets (9). Conceptually, alternative targets to CFTR aim to compensate for the abnormal dehydrated airway surface liquid that results from dysfunctional CFTR by modulating other ion channels, transporters and pumps (16-20) providing therapeutic options for those individuals with genotypes that do not produce CFTR protein and could assuage limited responses to existing CFTR modulators.

The clinical success rate of drugs in development is appreciably higher when there is human genetic evidence that supports a drug target (21). Genome-wide modifier gene studies in CF have aimed to identify genetic loci that impact disease severity in the presence of CFTR dysfunction in a hypothesis-free approach. First identified in a genome-wide association study (GWAS) of intestinal obstruction in CF (7), the C allele of rs7512462 in intron 5 of Solute Carrier Family 26 member 9 (*SLC26A9*) (chr1:205899595, hg19) has consistently demonstrated a beneficial effect for several CF co-morbidities, including in the exocrine (2, 3) and endocrine pancreas (4, 22).

These co-morbidities appear to originate from pre-natal dysfunction in the CF pancreas (1, 3, 23). According to the Genotype Tissue Expression project (GTEx; (24) and http://www.gtexportal.org/home/), rs7512462 is an expression quantitative locus (eQTL) for *SLC26A9* where the C allele is associated with greater expression in the adult pancreas (p=1.2×10^-5^, effect size=0.27). Rs7512462 and additional *SLC26A9* eQTLs in the region of chr1: 205,806,897- 206,006,897(hg19) colocalize with the CF co-morbidity (meconium ileus) GWAS summary statistics (1), supporting gene expression variation is responsible for the observed CF GWAS finding.

Given the importance of attenuating progressive lung disease in CF, there was significant interest in investigating the contribution of *SLC26A9* to lung function despite a lack of association evidence with rs7512462 across a broad CF population, including the largest CF GWAS of lung function to date (5). SLC26A9 is an anion chloride channel/transporter in epithelial cells that contributes to constitutive apical chloride conductance and enhances cAMP-regulated CFTR currents (25-28) and *Slc26a9*^-/-^/*Cftr^-/-^* mice show worse post weaning survival over *Cftr^-/-^* models (29). Rs7512462 appears to associate with both the residual and activated current in lung cells (30, 31). In individuals with *CFTR* variants that maintain apical membrane localization, rs7512462 showed evidence of association with CF lung function (31) and the relationship was greatly enhanced upon treatment with the CFTR modulator IVA where the C allele was associated with greater lung function improvement (31, 32). That rs7512462 was not shown to be a strong modifier of CF lung disease (5, 31) in the absence of treatment with CFTR correctors but is associated with lung function in a non-CF population study (9, 33) is consistent with functional studies, where differential SLC26A9 interaction with wild-type CFTR and with Phe508del-CFTR in human bronchial epithelia (HBE) has been reported (25, 31).

The functional and CF population studies highlight the potential of SLC26A9 as a target to improve CF outcomes across the affected organs and to improve response to approved CFTR modulators. However, several questions remain including whether population studies support SLC26A9 as providing alternative chloride transport in individuals with genotypes for which there are no approved therapies; whether enhancing SLC26A9 will improve lung function response to CFTR modulators targeting the most common Phe508del variant; which patient-specific cell-based models of the airway demonstrate SLC26A9 expression; and whether enhancing SLC26A9 could benefit other obstructive lung diseases such as chronic obstructive pulmonary disease (COPD) in light of the rs7512462 association with lung function in non-CF population studies (33).

Through continued recruitment into the Canadian CF Gene Modifier Study (CGMS) and in collaboration with the US PROSPECT observational study ((34); https://clinicaltrials.gov/ct2/show/NCT02477319), we investigate the association between rs7512462 and (1) lung function in individuals with CF with different *CFTR* genotypes including variants that result in no CFTR protein product (i.e. minimal function alleles); (2) individual lung function responses to CFTR modulators IVA and LUM/IVA; (3) CFTR function in primary cultured human nasal epithelia (HNE) from individuals with CF in response to approved and experimental CFTR modulators; and (4) other phenotypes in non-CF populations to inform the clinical utility of SLC26A9 modulators beyond applications in CF.

## Methods

### Study Design

The Canadian Cystic Fibrosis Gene Modifier Study (CGMS) is a registry-based observational genetics study. The US Part B Cystic Fibrosis Foundation (CFF) Therapeutics PROSPECT study is an observational study of modulator treatment. The current study investigates the relationship between rs7512462 and lung function pre- and post-CFTR modulators in Canadian and US CF cohorts; in the general population; and in a case control study of chronic obstructive pulmonary disease (COPD).

Clinical data collected as part of the CGMS include forced expiratory volume in 1 second (FEV_1_), age, sex, and height which are all obtained from the Canadian CF Patient Data Registry (CCFRD), with occasional augmentation by chart review at the participating sites. Genetic data is linked to clinical data through an approved data access agreement with Cystic Fibrosis Canada. Part B of the CFF Therapeutics PROSPECT observational study (https://clinicaltrials.gov/ct2/show/NCT02477319 and (34)) in the US evaluated the effectiveness of LUM/IVA and collected buffy coat and clinical data from individuals with two copies of the Phe508del variant who were prescribed LUM/IVA. We received the PROSPECT study buffy coat and corresponding clinical data from the US CFF. DNA was extracted for genotyping on Illumina platforms as described in Supplementary Information for sample genotyping and quality control.

### Phenotypes, Sampling and Inclusion Criteria

The CGMS has enrolled 3,257 participants from 9 provinces and 35 clinics across Canada with a range of *CFTR* genotypes that are reflective of the Canadian CF population. To investigate the association of *rs7512462* with pre-treatment lung function, we specifically study the subgroups of the CGMS who are: (1) homozygous for Phe508del; (2) carrying at least one copy of the G551D (c.1652G>A;p.Gly551Asp) variant or another gating variant approved for IVA; and (3) individuals with two *in trans* minimal function (MF) variants (https://www.cff.org/research-clinical-trials/types-cftr-mutations) for which there are no CFTR modulators approved; these include nonsense, splicing and small indel variants that cannot be targeted by CFTR modulators (Fig. 1). Lung function severity in the absence of modulator treatment for participants in CGMS was measured by Saknorm, the survival adjusted average CF-specific Kulich FEV_1_ percentiles that is normalized (5, 31, 35). For participants whose recruitment year was after 2008, Saknorm is calculated using Canadian CF-specific reference equations from 2008–2014 (36), rather than US reference equations (37). Saknorm enables comparison of CF lung disease across the age spectrum where there is differential mortality and is therefore used as a phenotype in CF genetic association studies. After quality control (QC), n=89 participants with at least one gating mutation, n=1,266 with homozygosity for Phe508del and n=63 with two MF alleles in trans are included in the analysis. Of these participants, 54 individuals with at least one gating mutation and 1,013 who are homozygous for Phe508del were also included in a previous publication (31) (Fig. 1).

**Fig.1.**
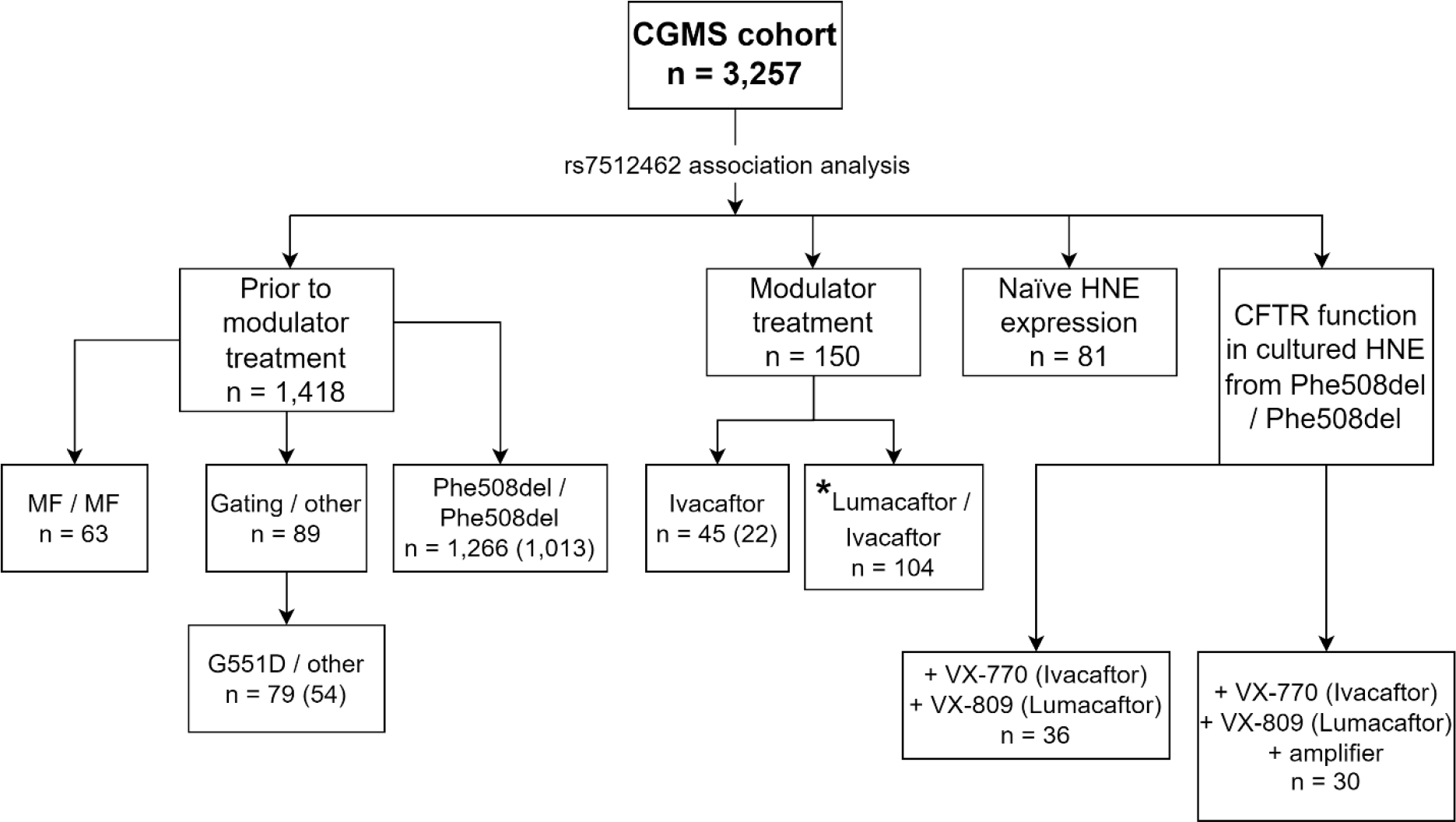
Assessment of the association of rs7512462 in the Canadian CF Gene Modifier Study (CGMS) population broken down according to particular sub-study groups. Sample sizes indicated in parentheses represent the overlap with those published in (31). *This analysis is combined with the US PROSPECT cohort.

CGMS participants prescribed IVA or LUM/IVA were also included in the treatment response study, together with the US PROSPECT participants for LUM/IVA (Fig. 1, Table 1). Participants from the PROSPECT study were younger and healthier than the Canadian cohort (as measured by forced expiratory volume in one second (FEV_1_) percent predicted (FEV_1pp_)) at treatment initiation (Table 1). Similar to (31), all participants included for modulator lung response analysis had a baseline measurement between 30 and 96 FEV_1pp_ measured within 3 months prior to, or on, the treatment initiation date. The CGMS data is obtained from the CCFRD with variable longitudinal entry depending on the resources of the individual clinics across the country while PROSPECT collected regular measurements at 1, 3, 6 and 12 months after treatment initiation. To account for this difference, the LUM/IVA treatment response is defined as the difference in FEV_1pp_ between the first visit on treatment within 5-7 months and that measured at baseline following convention in the literature (38); for the IVA study, aligned with reported treatment responses seen by 15 days (10), the difference between mean FEV_1pp_ within 15 to 400 days to FEV_1pp_ baseline (31), as well as the difference between the longitudinal FEV_1pp_ within 15 to 400 days to FEV_1pp_ baseline are used. We also investigated an IVA analysis with treatment response defined as the difference from the 1^st^ FEV_1pp_ within 15 to 60 days after the treatment initiation; the conclusion is comparable but because the sample size is reduced to 33, we report the mean treatment response and longitudinal treatment response in 15 to 400 days. After phenotype and genotype QC, 91 participants from PROSPECT and 104 CGMS participants were included in the genetic analysis for LUM/IVA and 45 for the IVA study (see Supplementary Table 1 for sample exclusion; Fig. 1).

**Table 1.** Characteristics of participants included for response to CFTR modulators from the CGMS and US PROSPECT studies. The lung function measure used for the CFTR modulator study is forced expiratory volume in one second percent predicted (FEV_1pp_). A subset of n=22 individuals from CGMS on IVA were included for analysis in (31) while n=23 are newly- recruited into the CGMS.

| CGMS Participants in CFTR modulator study |  |  |  |  | US PROSPECT |
| --- | --- | --- | --- | --- | --- |
| Participants on CFTR modulator | Participant Characteristics | CGMS IVA (In (31), n=22<sup>*</sup>) | CGMS IVA (new, n=23) | CGMS LUM/IVA (n=104) | LUM/IVA (n=91) |
| rs7512462 | TT/TC/CC | 11/10/1 | 12/9/2 | 41/48/15 | 30/50/11 |
| age | mean (range) | 26.49 (6.1-58.3) | 31.3 (14.0-55.3) | 25.7 (10.5-55) | 20.9 (6-57) |
|  | age ≤ 12 | 5 | 0 | 2 | 23 |
|  | age (12,20] | 4 | 3 | 38 | 27 |
|  | age (20,30] | 3 | 11 | 38 | 24 |
|  | age>30 | 10 | 9 | 26 | 17 |
| sex | female (%) | 16 (72.7%) | 13 (56.3%) | 56 (53.8%) | 52 (57.1%) |
| FEV<sub>1pp</sub> baseline | mean (range) | 66.6 (30.7-90.4) | 58.9 (30.9-89.6) | 59.6 (30.8-95.5) | 75.7 (31.7-95.8) |
| Number of post-treatment FEV<sub>1pp</sub> in 400 days | median (range) | 4 (1-6) | 4 (1-20) | 6 (1-33) | 4 (2-4) |

<sup>\*</sup> one sample with FEV<sub>1pp</sub> baseline outside the inclusion criteria and another who received a lung transplant were removed.

We also investigated CFTR function in 46 CF Canada SickKids Program in CF Individualized Therapy (CFIT; (39)) participants homozygous for Phe508del whose nasal epithelia were brushed, cultured to passage 2 (P2) and mounted in a circulating Ussing chamber. Thirty-six individuals who underwent brushing prior to modulator initiation and for whom we had rs7512462 genotype are included in the Ussing chamber analysis (Supplementary Information for additional cell culture and Ussing chamber study information; Fig. 1).

A subset of the CGMS cohort (n=82; Fig. 1) and 6 healthy controls had RNA from their nasal cells sequenced as part of CFIT. In addition, we have sequenced nasal cells for 9 CFIT samples cultured to passage 3 (P3) at two time points (14-16 days and 26-30 days). We also sequenced the RNA from de-identified CF individuals who underwent lung transplantation, obtained from paired human bronchial and nasal epithelia; n=17 independent pairs of uncultured primary HNE and HBE and n=16 cultured HNE and HBE pairs (Supplementary Information for sampling and cell culturing).

Illumina Smart-Seq2 single-cell RNA data from three individuals were obtained from the Human Lung Cell Atlas (40). The donors consisted of two males and one female, aged between 46 to 75 years. The samples were freshly resected lung tissues obtained during surgery with confirmed normal histology (except for very mild emphysema in some of the samples from one individual).

Summary association statistics from other GWAS phenotypes were obtained from the GWAS ATLAS (41) (Supplementary Information for PheWAS data extraction). Association between rs7512462 and lung function was assessed using the UK Biobank data under application #40946, or was investigated using summary statistics from (33) when available.

### Association Analysis of rs7512462

#### i Association with lung function (Saknorm) by CFTR genotype group

To assess whether rs7512462 was associated with Saknorm in the different *CFTR* genotype groups, we carried out a stratified analysis, separately for those with gating variants (or the subset specifically with G551D), those homozygous for Phe508del and those with two minimal function (MF) variants, using both additive and recessive models. In each CFTR genotype subgroup, the association analysis used the R package „rms’ (42) to obtain a robust variance estimator and the linear regression included an indicator for which reference cohort was used to calculate the Saknorm phenotype. For individuals with at least one G551D variant, the rs7512462 association in CGMS is meta-combined with that from four previously published cohorts: a cohort from France (32) and 3 from the United States (43). For CF individuals homozygous for Phe508del or for those with at least one gating mutation, the rs7512462 association results are also meta-combined with those reported in (32). Meta-analysis is implemented using inverse variance and sample size weighting with the R functions „metagen’ in the package „meta’ (44) and „rma’ in the package „metafor’ (45), respectively and the R package „forestplot’ is used to generate forest plots (46). To assess whether lung function is different between the three *CFTR* genotype groups, we regressed Saknorm on two indicators for the three CFTR genotype categories. Additionally, to assess whether the effects of rs7512462 genotype on Saknorm were different between the three *CFTR* genotype groups, we included an SNP*-CFTR* interaction term in the regression model and performed an F-test using the aov function. All analyses adjusted for the reference cohort used to calculate Saknorm and only two-sided p-values less than 0.05, and with the direction that the C alleles is beneficial, are considered significant.

#### ii Association with response to modulator treatment

We also use multivariable regression with robust variance estimation to assess the association of rs7512462 with the modulator FEV_1pp_ response, where rs7512462 is coded recessively to align with what is observed in exploratory data analysis (Fig. 2). For the association with treatment response to IVA, covariates include age and FEV_1pp_ at baseline. To account for the variable follow-up days and the variable baseline measure time prior to the treatment initiation, we also implement a linear mixed-effect model for participants on IVA and with follow-up measurements between 15 to 400 days using the R function „lmer’ in the package „lme4’ with a random intercept (47).

**Fig. 2.**
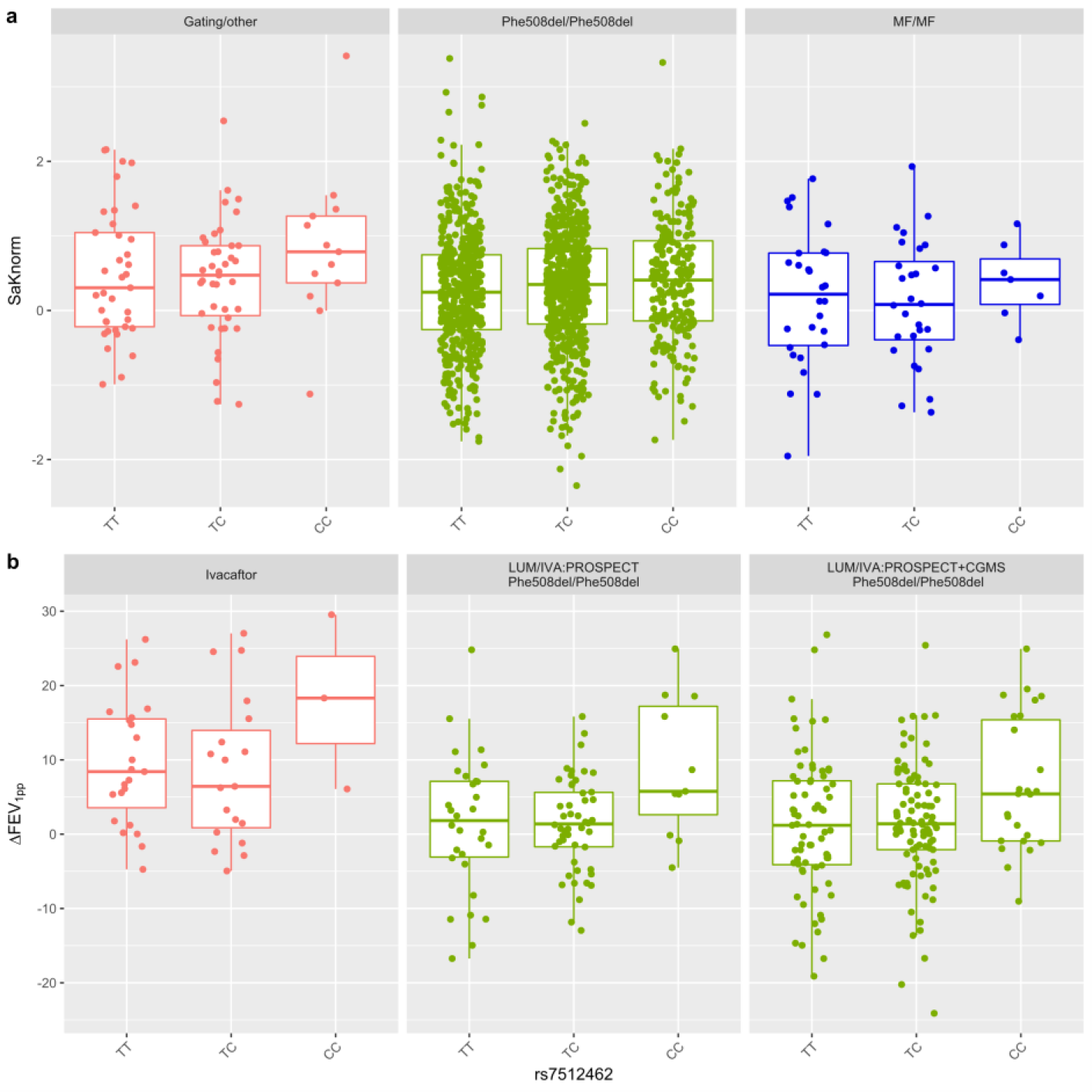
Participant lung function or lung treatment response categorized by rs7512462 genotype. (a) Boxplots of lung function from CGMS participants prior to modulator treatment, measured as Saknorm across different CFTR mutation groups. Saknorm is calculated as in (5, 35) with FEV_1_ measurements taken prior to modulator treatment, if applicable. (b) Boxplot includes an overlay with a stripchart for treatment response in patients on IVA (n=45), LUM/IVA in PROSPECT cohort (n=91) or LUM/IVA in combined PROSPECT and CGMS samples (n=195). The center line, upper and lower box limits of boxplots correspond to the median, the third quartile (Q3) and the first quartiles (Q1), respectively; whiskers reflect the 1.5x interquartile range (IQR), i.e. bottom whisker is Q1-1.5xIQR and upper whisker is Q3 +1.5 x IQR). Each dot represents an individual measurement. Following (31), the treatment response for IVA is defined as the difference between the mean post-treatment FEV_1pp_ within 15 to 400 days and FEV_1pp_ baseline. Response for LUM/IVA is defined as the difference in FEV_1pp_ between the first post treatment within 5 to 7 months to the baseline ((38); Methods).

For treatment response to LUM/IVA, besides rs7512462, age and FEV_1pp_ at baseline, principal components (PCs) were also included to adjust for population structure, which were calculated from the PROSPECT or the combined CGMS and PROSPECT studies by the R function PC-AiR in the GENESIS package (48, 49) using the kinship matrix estimated by KING 2.2.4 (50). The significant PCs were selected based on the Tracy-Widom test using the function twtable in POPGEN of Eigensoft (51) ; 7 PCs were included for the US PROSPECT analysis and 4 PCs for the US PROSPECT+CGMS combined analysis. Analysis of the Ussing chamber data to assess association of CFTR functional response to CFTR modulators with rs7512462 uses multivariable regression with a robust variance estimator, adjusted for a binary indicator of culture media. The boxplots are obtained using the functions ggplot and geom_boxplot from the package ggplot2 in R (52) and the function geom_jitter from ggplot2 is used to overlay the individual measurements in a stripchart.

#### iii Association analysis with COPD and non-CF lung function in the UK Biobank and SpiroMeta Consortium

Genetic association analyses for spirometry measures in the UK Biobank were conducted using imputed (v3) phenotypic data obtained from the UK Biobank (53, 54). In-house scripts, R package rbgen and C++ tool bgenix (55), were used to index, subset and perform association analysis using imputed dosage (53). COPD was defined according to the GOLD (levels 2 to 4) criteria of moderate to very severe airflow limitation (56) (FEV_1_/FVC ratio < 0.7 and FEV_1pp_ < 80%). Prior to analysis, spirometry and genotyping quality control (QC) were carried out as suggested in (33), with the exception of kinship and ethnicity analyses, where we opted to use KING’s (v.2.0) --unrelated option (50) and the UKBB’s UID 22006 for the identification of Europeans. This QC method is more conservative and yielded 263,461 participants compared to 321,047 participants in (33). We removed individuals with incomplete data for sex (UID=22001), age (UID=21022), height (UID=50), and smoking status (ever/never, UID=20160). FEV_1pp_ was calculated using the Global Lungs Initiative (GLI) calculator using the best FEV_1_, age (UID=21022), height (UID=50), and sex (UID=22001). From these, 22,071 participants fit the spirometrically-defined COPD criteria. Summary statistics for population-level GWAS of spirometry (FEV_1_/FVC ratio, and peak expiratory flow (PEF)) were obtained from meta-analysis of the UKBB and SpiroMeta Consortium (33). Phenotypic analysis at the *SLC26A9* locus included the best measures for PEF, FEV_1pp_ and FEV_1_/FVC ratio. The best measure for FEV_1_ (UID=20150) and FVC (UID=20151) were defined as the highest measure from the array of values of up to three blows (UID=3063 and 3062, respectively) that were deemed acceptable according to blow quality metrics (UID=20031): “blank”, “ACCEPT”, “BELOW6SEC ACCEPT” and “BELOW6SEC”; and a back-extrapolated volume (derived from the blow curve time series, UID=3066) greater than 5% or less than 150mL, as explained in (33). The best FEV_1_/FVC is calculated from the selected best FEV_1_ and FVC after determining blow reproducibility within 250mL from any other blow as explained in (33). Principal components were calculated using flashPCA2 v2.1 (57) which is designed for Biobank-scale data. All association models included three principal components, sex, age, age^2^, sex × age, sex × age^2^ and smoking (ever/never). All spirometry measures were inverse rank normal transformed prior to association analysis using the RNOmni R package’s (v0.7.1) rankNorm function (58). Association summary statistics from each of the lung function phenotypes were then used for colocalization analysis in LocusFocus (v1.4.9; (59)).

### Haplotype analysis of CGMS in CF participants homozygous for Phe508del

Risk haplotypes that include rs7512462 were reported to associate with age-of-onset of CF-related diabetes in individuals homozygous for Phe508del (60) and here we similarly construct these haplotypes and assess their association with Saknorm. Lam et al (60) constructed the haplotypes from 41 common SNPs (MAF>15%) in the same linkage disequilibrium (LD) block as rs7512462 and found that two common haplotypes are associated with later CFRD onset (low risk (LR), minor haplotype frequency [MHF]=28.4%, p=1.14×10^-3^) and earlier CFRD onset (high risk (HR), MHF=24.1%, p=4.34×10^-3^) (n=594) when using PLINK’s haplotype analysis command (--chap and --each-vs-others).

To mimic this haplotype analysis we used imputed genotype data from the CGMS (1) in individuals homozygous for Phe508del (n=1,266). The imputed data included 40 of the 41 variants used by Lam et al (60), with the multi-allelic variant rs144469431 removed. A subset of n = 447 CGMS participants sequenced using 10X Genomics linked-read whole genome sequencing technology (10XG, Supplementary Information), which facilitates improved phasing, was used as the reference panel to phase these imputed data into haplotypes. We then implemented two analyses. The first analysis used the same PLINK command as in (60) in an unrelated set of CGMS Phe508del/Phe508del participants (n=1,164). The second analysis is in a larger subset that includes related individuals and conducts haplotype association using linear regression with a robust variance estimate that accounts for the relatedness (n=1,266), comparing each of the eight haplotypes studied in (60) to a comparison group that includes all others using an additive model.

### RNA Sequencing, Quality Control and analysis

The CF human nasal epithelia (HNE) cells were sequenced on the Illumina HiSeq 2500 platform (Illumina Inc. San Diego, California, USA) and aligned as described in (39). Expression counts were quantified using RNA-SeQC (ver. 2.0.0) and normalized to transcripts per million (TPM) (61) as well as trimmed mean of M values (TMM) measures (62) .

To compare the expression level for *SLC26A9* across the different airway models, we calculated the TPM from naïve HNE for 82 CGMS participants and 6 healthy controls, 16 pairs of de-identified cultured HNE and HBE, 17 pairs of de-identified naïve HNE and HBE and 9 CFIT cultured HNE samples.

eQTLs were calculated from 79 CGMS participants for whom both genotype and RNA sequence from naïve HNEs were available (Fig 1). Quality control required TPM ≥ 0.1 and read counts ≥ 6 in greater than 20% of the sample to be analyzed. FastQTL (ver. 2.0) was used to conduct differential gene expression analysis of TMM-normalized read counts on SNP genotypes (63). Covariates included the top 3 genotype principal components, 15 probabilistic estimation of expression residual (PEER) factors, sample study source, sex, genotyping platform, RNA integrity number (RIN) and PTPRC/CD45 gene expression which adjusts for immune cell composition in the samples. The genotype principal components and PEER factors were generated using R packages GENESIS (48, 49) and peer (64), respectively. The average expression level of *SLC26A9* was compared across HBE and HNE in both cultured and naïve paired tissues. Paired t-tests were conducted using the *t.test()* function in R v3.6.1 (65), based on TPM.

For the Human Lung Atlas single cell data (40), cellular expression profiles were clustered by nearest-neighbor for visualization. RNA count data and cell annotations provided in this dataset were used; counts were normalized (library-size corrected) to 1,000,000 reads per cell and log-scaled. Scanpy v1.8.2 in Python 3.7 was used for the *CFTR* and *SLC26A9* single-cell expression visualization of the Human Lung Cell Atlas data (66).

A subset of these samples with expression from more than 500 genes and greater than 50,000 total mapped reads were used for statistical modeling of CFTR-SLC26A9 co-expression. Library sizes were normalized using the TMM method. Log-transformed read counts per million (log- CPMs) were generated using the normalized library sizes and were used to calculate the Spearman’s correlation between the *CFTR* and the *SLC26A9* genes. Expression raw read count from *CFTR* was regressed on that of *SLC26A9* in the zero-inflated negative binomial model analysis with a log link function. Normalized library size was adjusted using an offset term. Zero-inflation was modelled by the detection rate variable which is the proportion of genes with expression data in a cell. The effect size estimate indicates log-ratio relations between *CFTR* and *SLC26A9* expression. The Spearman’s correlation and the zero-inflated negative binomial model analysis were performed by R function cor.test() and package pscl (67), respectively.

## Results

### rs7512462 is associated with CF lung function in the absence of treatment measured as Saknorm

Using the International CF Gene Modifier consortium lung phenotype, Saknorm ((35); Methods), we compared lung function across the three *CFTR* genotype groups. Saknorm measures differ significantly between individuals with a gating variant (n=89) versus those who are homozygous for Phe508del (n= 1,266; effect size=0.20, p=0.030), while individuals with two MF variants (n=63) do not differ in Saknorm from Phe508del homozygotes (effect size= -0.11, p=0.27).

In CGMS participants with at least one G551D variant (n=79), rs7512462 did not reach statistical significance at the 0.05 level with Saknorm (effect size = 0.185, p=0.225). Meta- combining the CGMS association evidence with association results from four independent cohorts, a cohort from the French gene modifier study (FGS) with n=49 participants (32) and three cohorts with n=272 participants reported in Eastman et al (43), results in an effect size of 0.11 with p=0. 05 and an effect size=0.12 with p=0.036 from inverse variance weighted and from sample size weighted meta-analyses, respectively (Fig. S1; Supplementary Table 2). Meta-analysis of rs7512462 Saknorm association from the FGS (32) and CGMS cohorts with at least one gating variant approved for ivacaftor have similar but more variable effect size than that from the G551D subset (effect size =0.12 with p=0.18; Supplementary Table 2).

In individuals from the CGMS who are homozygous Phe508del (n=1,266), each C allele is associated with an increase in lung function, albeit with a smaller effect size (effect size = 0.069, p=0.028, Fig. 2a; Fig. S2). Meta-analysis of this CGMS association with an rs7512462 Saknorm association in n=1,802 individuals who are Phe508del/Phe508del from the FGS (32) also show association in both inverse variance weighted (effect size=0.050, p=0.021) and sample size weighted (effect size=0.046, p=0.037) meta-analyses (Fig. S2; Supplementary Table 2). Constructing haplotypes in the CGMS in individuals homozygous for Phe508del as in (60) does not provide evidence for significant low risk (LR) or high risk (HR) haplotypes contributing to Saknorm variation (Supplementary Table 3) and does not provide greater power than using the single SNP rs7512462 to mark the locus for association with lung function (Supplementary Result; Supplementary Table 3).

Importantly, in individuals with two MF variants, the CC genotype is associated with improved lung function when compared to individuals with the TT and TC genotype (effect size=0.45, p=0.0083, Fig. 2a). This is consistent with the hypothesis that for rs7512462 marking SLC26A9 activity, SLC26A9 may be providing alternative chloride transport properties in individuals with *CFTR* variants for whom no current approved therapies exist.

When combining CGMS participants from the three genotype groups in a joint multivariable regression model to assess the association with rs7512462, the C allele is associated with greater lung function (p=0.017 and 0.022 for additive and recessive models, respectively, Table 2). Fitting an interaction term in this multivariable model, the C allele does not demonstrate a statistically significant difference in effect size depending on one’s *CFTR* genotype (interaction p-values 0.90 and 0.46 for additive and recessive models, respectively).

**Table 2.**
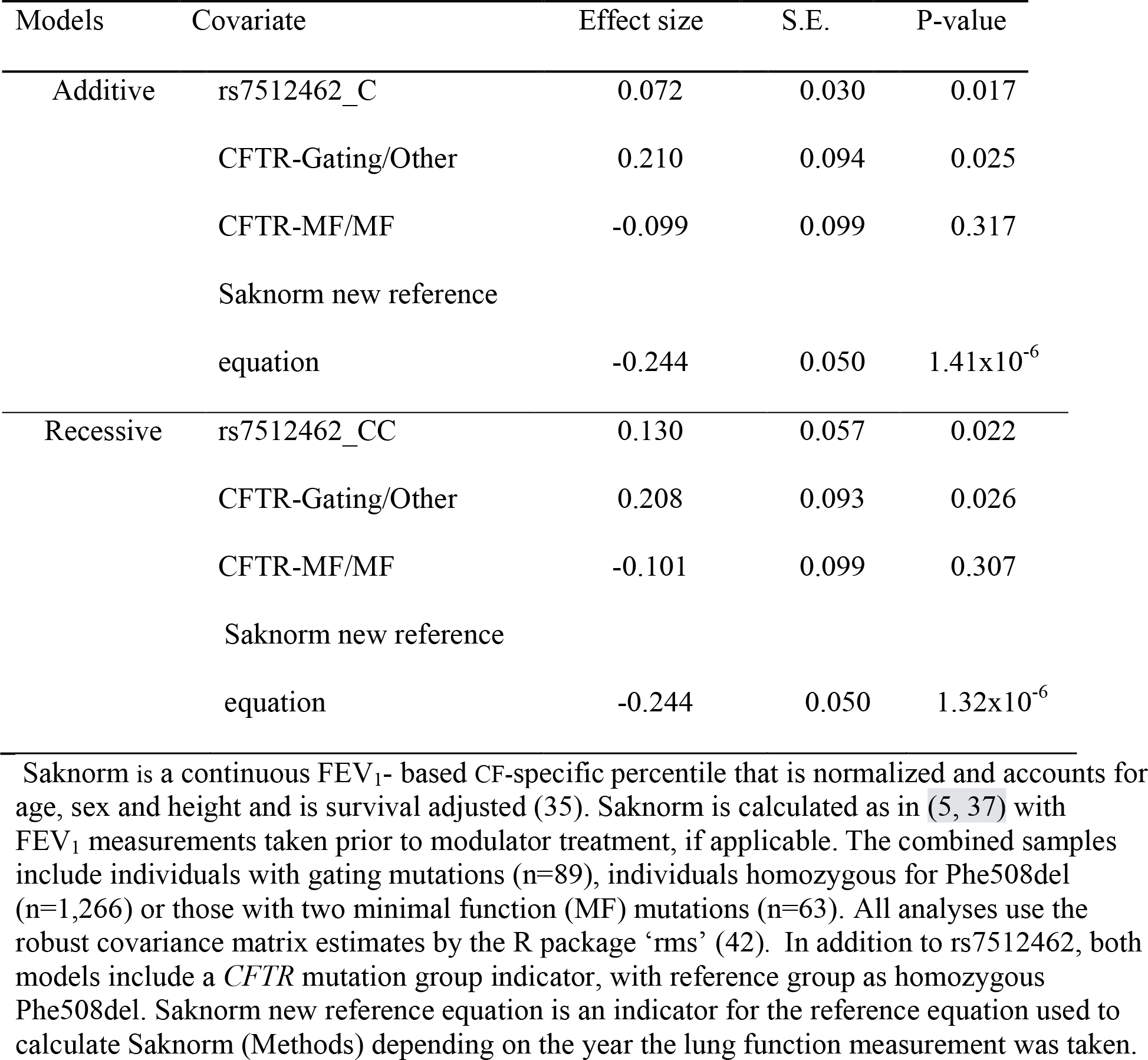
Association between Saknorm and rs7512462 using additive and recessive codings in the CGMS cohort including individuals with a gating variant, individuals who are homozygous Phe508del or those who carry two MF alleles.

| Models | Covariate | Effect size | S.E. | P-value |
| --- | --- | --- | --- | --- |
| Additive | rs7512462_C | 0.072 | 0.030 | 0.017 |
|  | CFTR-Gating/Other | 0.210 | 0.094 | 0.025 |
|  | CFTR-MF/MF | -0.099 | 0.099 | 0.317 |
|  | Saknorm new reference equation | -0.244 | 0.050 | 1.41x10<sup>-6</sup> |
| Recessive | rs7512462_CC | 0.130 | 0.057 | 0.022 |
|  | CFTR-Gating/Other | 0.208 | 0.093 | 0.026 |
|  | CFTR-MF/MF | -0.101 | 0.099 | 0.307 |
|  | Saknorm new reference equation | -0.244 | 0.050 | 1.32x10<sup>-6</sup> |

Saknorm is a continuous FEV<sub>1</sub>- based CF-specific percentile that is normalized and accounts for age, sex and height and is survival adjusted (35). Saknorm is calculated as in (5, 37) with FEV<sub>1</sub> measurements taken prior to modulator treatment, if applicable. The combined samples include individuals with gating mutations (n=89), individuals homozygous for Phe508del (n=1,266) or those with two minimal function (MF) mutations (n=63). All analyses use the robust covariance matrix estimates by the R package ‘rms’ (42). In addition to rs7512462, both models include a *CFTR* mutation group indicator, with reference group as homozygous Phe508del. Saknorm new reference equation is an indicator for the reference equation used to calculate Saknorm (Methods) depending on the year the lung function measurement was taken.

### Rs7512462 C allele is associated with improved response to CFTR modulators

Newly recruited participants into the CGMS on IVA (n=23) were, on average, older and had worse baseline lung function than those included in (31) (Table 1). Despite the difference in baseline characteristics and disease severity, we do observe additional supportive evidence for improved lung function response when combining the newly and previously recruited participants (difference in FEV_1pp_ pre and post treatment initiation) in individuals with the CC genotype upon treatment with IVA (effect size=9.9, p=0.03, Table 3; Fig. 2b). Results from a linear mixed-effect model which accounts for the longitudinal nature of the data provides consistent results (effect size =12.76, p=0.025; Supplementary Table 4).

**Table 3.** Association of rs7512462 for CFTR modulator response.

| Studies | Covariates | Effect Size | S.E. | P-value |
| --- | --- | --- | --- | --- |
| IVA(CGMS) | Age at baseline | -0.077 | 0.095 | 0.419 |
|  | FEV<sub>1pp</sub> at baseline | -0.025 | 0.073 | 0.731 |
|  | rs7512462_CC | 9.905 | 4.477 | 0.033 |
|  | Early CGMS Cohort | 5.336 | 2.606 | 0.047 |
| LUM/IVA (US PROSPECT) | Age at baseline | -0.188 | 0.074 | 0.014 |
|  | FEV<sub>1pp</sub> at baseline | -0.123 | 0.053 | 0.024 |
|  | rs7512462_CC | 8.518 | 2.657 | 0.002 |
| LUM/IVA (US PROSPECT+CGMS) | Age at baseline | -0.119 | 0.064 | 0.066 |
|  | FEV<sub>1pp</sub> at baseline | -0.069 | 0.037 | 0.066 |
|  | rs7512462_CC | 5.217 | 1.871 | 0.006 |
|  | PROSPECT Cohort | 0.865 | 1.401 | 0.540 |

CFTR modulator response is the difference between the FEV<sub>1pp</sub> after the modulator response and the value at last visit before the treatment initiation within 3 months as defined in Methods. Increased $\Delta$ FEV<sub>1pp</sub> indicates better treatment response. Sample sizes in each CFTR modulator treatment group are n=45 for IVA (CGMS), n=91 for LUM/IVA (US PROSPECT) and n=195 for the combined LUM/IVA analysis of the US PROSPECT and CGMS participants (US PROSPECT+CGMS). Early CGMS Cohort for IVA-CGMS reflects an indicator for those participants that were included in an earlier published study (31). LUM/IVA studies were also adjusted for population structure by PCs (n=7 PCs for US PROSPECT and n=4 for US PROSPECT+CGMS). PROSPECT Cohort is an indicator of whether the individual was a participant in the US PROSPECT study or the CGMS. All results use robust variance estimates.

Through ongoing recruitment in the CGMS there were 104 participants prescribed LUM/IVA clinically. In collaboration with the US PROSPECT observational study, we investigated lung function response to LUM/IVA alone (n=91) and in a combined sample of n=195 individuals homozygous for Phe508del stratified by rs7512462. Despite minimal clinical response to LUM/IVA reported on average (14), we do observe a significant improvement in lung function response in those with the CC genotype (p=0.002 in US PROSPECT and p= 0.006 for combined, respectively; Table 3), akin to observations for IVA (Fig. 2b and (31)), and in studies of improved CFTR function with the rs7512462 C allele in HNE (30) and HBE (18, 31) cells obtained from individuals homozygous for Phe508del. Thus, the rs7512462 genotype shows improved response to the LUM/IVA combination therapy in cohorts of CF patients who were monitored observationally during their real-world experience with the approved modulators.

We investigated CFTR-mediated current in 36 cultured HNE from CGMS participants homozygous for Phe508del upon treatment with VX-770+VX-809 (as in (30), corresponding to the IVA/LUM combination; VX-770 applied acutely) and upon treatment with VX-770+VX-809 + an amplifier under experimental investigation ((68); Fig. 3). We used the HNE from the earliest passage available to us (P2) to align with (30). Defining the treatment response as the difference in ΔIeq -forskolin from DMSO to VX-770+ VX809 (n=36) or VX-770+ VX-809 + amplifier (n=30), we see a significant improvement in CFTR function in the CC group (effect size=-1.934, p=0.0022, Table 4) in HNE with VX-770+VX-809+ amplifier. The increase in CFTR function in the CC group in the cultured HNE with VX-809 does not reach statistical significance (effect size=-0.406, p=0.338, Table 4) unlike previous reports in HNE and in HBE models, although the direction of effect is consistent. This difference in effect size made us question the *SLC26A9* gene expression profile across different lung tissue models, especially given the low passage of the primary HNE cultured cells studied by (30).

**Fig.3.**
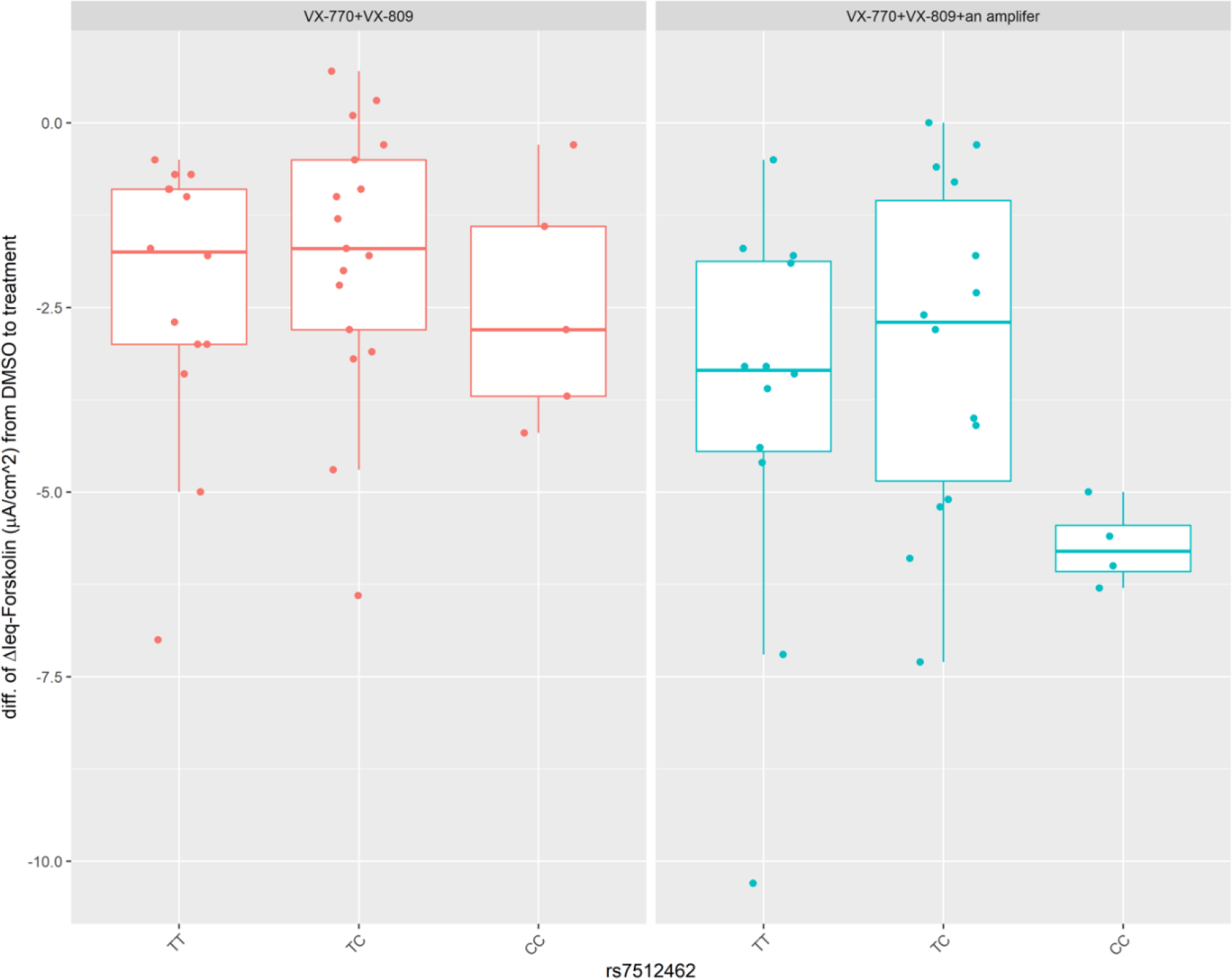
Boxplot overlayed with stripchart for CFTR function by rs7512462 genotype in HNE cultured to P2. The center line, upper and lower box limits of boxplots correspond to the median, the third quartile (Q3) and the first quartiles (Q1), respectively; whiskers reflect the 1.5x interquartile range (IQR), i.e. bottom whisker is Q1-1.5xIQR and upper whisker is Q3 +1.5 x IQR). Each dot represents an individual measurement. The CFTR function is defined as the difference in ΔIeq-forskolin from treatment with VX-770+VX-809 (left; n=36) or VX-770+VX- 809+amplifier (right; n=30) to DMSO vehicle in CFTR-mediated current in cultured HNE from participants homozygous for Phe508del. More negative measurements reflect greater CFTR function.

**Table 4.** Association of rs7512462 for change of function to modulator treatment.

| Studies | Covariates | Effect Size | S.E. | P-value |
| --- | --- | --- | --- | --- |
| CFTR-mediated current (VX-770+VX-809) | rs7512462_CC | -0.406 | 0.417 | 0.3375 |
|  | Standard media | -1.969 | 0.456 | 0.0001 |
| CFTR-mediated current (VX-770+VX-809+ amplifier) | rs7512462_CC | -1.934 | 0.572 | 0.0022 |
|  | Standard media | -2.170 | 0.697 | 0.0044 |

Forskolin-stimulated currents mediated by CFTR (measured as change in current after application of forskolin, $\Delta I_{eq}$ -forskolin) is used as the treatment response; more negative values correspond to more CFTR activity. Functional study includes HNEs from individuals homozygous for Phe508del with VX-770+VX-809 (n=36) and VX-770+VX-809+ amplifier (n=30; a subset of the 36 samples tested with 770+VX-809) measured as difference in CFTR-mediated current after applying forskolin regressed on rs7512462 with a recessive coding. All results use robust variance estimates.

### SLC26A9 expression differs across airway models

While investigating eQTLs in various airway tissue models, we observed that there is no evidence supporting rs7512462 as an eQTL in the lungs from GTEx v8 (p=0.71) or from RNA obtained from naïve HNE of individuals with CF (p=0.64, n=79) despite its association with residual and activated current in Ussing chamber studies (30, 31). *SLC26A9* expression appears generally low across several different lung model systems we investigated, with an average of 1.34 transcripts per million (TPM) (Fig. S3a). We generated a resource that contains the transcriptomes from paired cultured and fresh naïve HNE and HBE tissue on the same individuals (Methods; GEO ID: GSE172232)). Of the primary lung cell models we investigated, the greatest expression is in the naïve HBE cells (TPM=1.94; Fig. S3a), and this expression level is 2.1-fold greater than in the naïve HNE (p=0.04, paired analysis in n=17 HNE-HBE naïve pairs). Unfortunately, naïve HBE cell models are not generally accessible for personalized medicine approaches (39), and cultured models are the norm. Here cultured HBE show mean expression with TPM=1.71 that is 2.5-fold greater than the CF cultured HNE (p=0.02, paired analysis for n=16 HNE-HBE cultured pairs), although there is some indication that a reduction in culturing time results in greater *SLC26A9* expression in the HNE (Fig. S3b).

Using single-cell RNA-seq data catalogued in the Human Lung Cell Atlas (40), we investigated the expression of *SLC26A9* and *CFTR* across cell types. Within lung cells, both genes are expressed in the epithelial cells of the alveoli and airway, particularly within alveolar type 2 (AT2) cells and club cells (Fig. 4). These cell types are also supported by single-cell Human Protein Atlas data ((69); http://www.proteinatlas.org/), which reports expression in AT2 cells (normalized TPM of 50.1 for *SLC26A9* and 65.6 for *CFTR*) and club cells (normalized TPM of 2.7 for *SLC26A9* and 17.8 for *CFTR*). Furthermore, single-cell GTEx data ((70, 71); http://www.gtexportal.org/home) shows concordant evidence where, *SLC26A9* and *CFTR* reads are observed in an appreciable fraction of AT2 cells (24.59% CFTR, 12.41% SLC26A9) and club cells (28.79% *CFTR*, 5.45% *SLC26A9*).

**Fig. 4.**
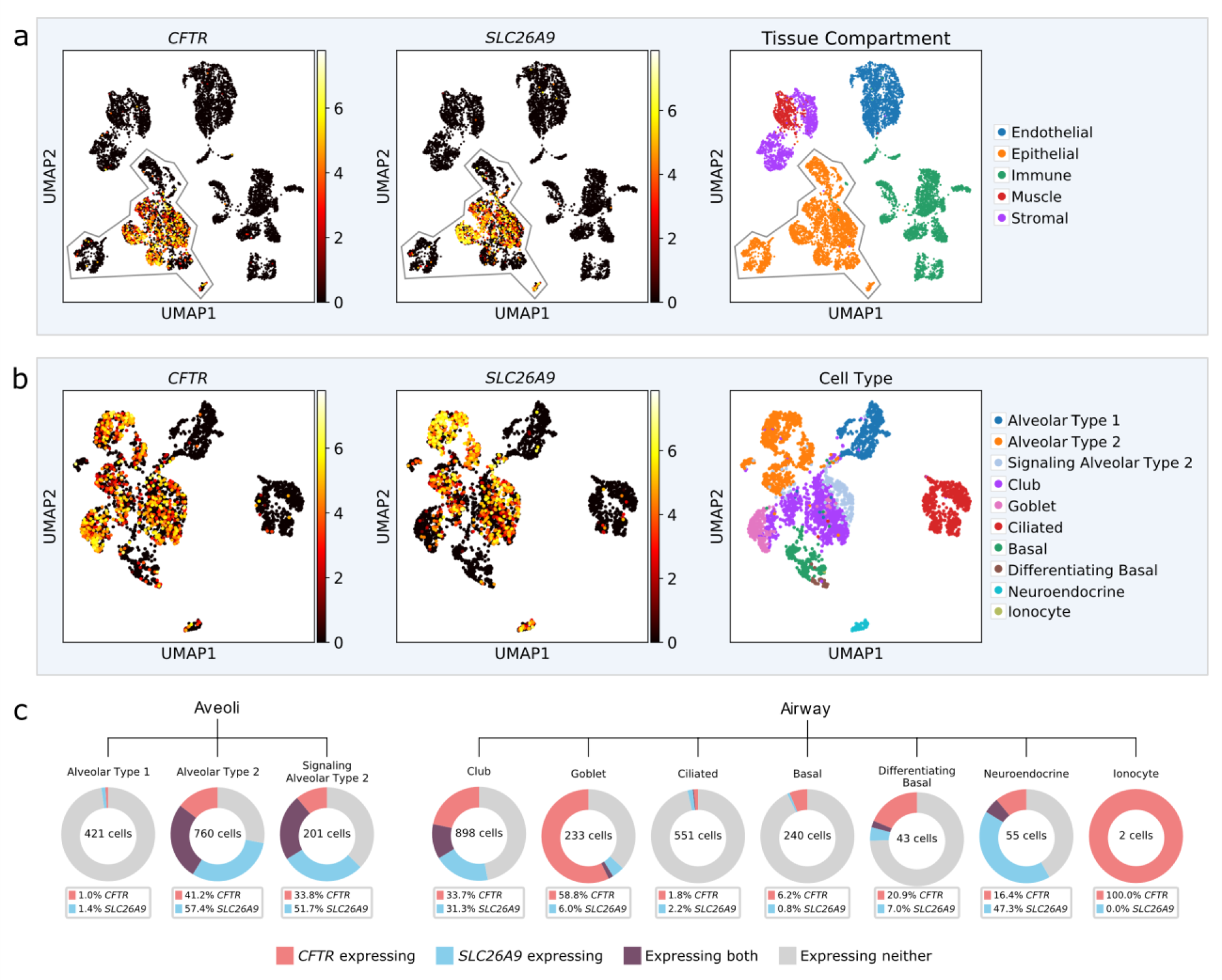
CFTR and SLC26A9 single-cell expression from the Human Lung Cell Atlas (40). (a) Lung cell expression profiles from three individuals were clustered by nearest-neighbor and visualized using uniform manifold approximation and projection (UMAP). Normalized and log- scaled RNA counts for CFTR and SLC26A9 are plotted for each cell. Tissue compartment as defined by the Human Lung Cell Atlas is shown; expression of both CTFR and SLC26A9 is primarily restricted to epithelial cells; (b) UMAP clustering of epithelial lung cells only, cell types were annotated in the Human Lung Cell Atlas and (c) Breakdown of CFTR and SLC26A9 expression by cell type. Cells were grouped as having CFTR read count > 1 (red), SLC26A9 > 1 (blue) or co-expressing both (purple). Cell total varies and is annotated for each cell type.

We assessed the evidence for *CFTR*-*SLC26A9* co-expression among the club, basal and alveolar epithelial cell types identified in the Human Lung Atlas study (40). Both the Spearman’s correlation and the zero-inflated negative binomial model indicated significant association between the *CFTR* and the *SLC26A9* read counts for AT2 cells (Table 5). The model estimate suggests positive co-expression between the two genes (log ratio = 0.00035, p=0.00974).

**Table 5:** Co-expression between *SLC26A9* and *CFTR* using the Human Lung Atlas data (40).

| Cell Type | Number of Cells | Zero-Inflated Negative Binomial Model |  |  | Spearman's Correlation |  |
| --- | --- | --- | --- | --- | --- | --- |
|  |  | Effect Size | S.E. | P-Value | Rho | P-Value |
| Club | 895 | -0.00007 | 0.00125 | 0.9564 | 0.05173 | 0.12196 |
| AT2* | 760 | 0.00035 | 0.00013 | 0.0097 | 0.13215 | 0.00026 |
| Basal | 240 | -11.65287 | 94.65422 | 0.9020 | -0.02365 | 0.71544 |
| AT1* | 421 | -8.41119 | 152.8630 | 0.9561 | -0.01178 | 0.80962 |
| AT2-s* | 201 | 0.00143 | 0.00122 | 0.2433 | 0.28686 | 0.00004 |
| Differentiating Basal | 43 | -0.02385 | 0.01931 | 0.2168 | 0.07235 | 0.64478 |
| AT2 and AT2-s | 961 | 0.00041 | 0.00014 | 0.0036 | 0.16567 | < 0.00001 |
| AT1, AT2 and AT2-s | 1382 | 0.00048 | 0.00015 | 0.0018 | 0.32261 | < 0.00001 |

The effect size estimate from zero-inflated negative binomial model indicates log-ratio relations between *CFTR* and *SLC26A9* expression. Values greater than zero correspond to positive co-expression. Spearman's correlation estimates (rho) were calculated using log-transformed read counts per million measures for *CFTR* and *SLC26A9*. The Spearman's correlation and the zero-inflated negative binomial model analysis were performed by R function cor.test() and package pscl (67), respectively. All p-values indicate two-sided statistical test results.
\*: AT2: Alveolar Epithelial Type 2; AT1: Alveolar Epithelial Type 1; AT2-s: Signaling Alveolar Epithelial Type 2

### Phenome-wide Association Study (PheWAS) of rs7512462 and colocalization analysis

We used the GWAS ATLAS database (41) that includes 4,756 GWAS from 473 unique studies with 3,302 unique traits, and the UK Biobank resource to investigate other non-CF traits associated with rs7512462. The 10 traits with the smallest reported p-values are listed in Table 6, four of which are respiratory phenotypes from the UK Biobank and Spirometa consortium (33): PEF and FEV_1_/FVC ratio. Saknorm, the lung function measurement used in the CF GWAS, is also calculated from FEV_1_ (5, 35). The list of significant phenotypes also includes our own CF modifier gene study where we first identified rs7512462 as a modifier of intestinal obstruction at birth, meconium ileus with evidence of an exocrine pancreatic origin ((1); Table 6). Interestingly, an earlier age at menarche (which is associated with weight) and a higher male waist circumference and waist-hip ratio are also associated with the rs7512462 C allele. The association of rs7512462 with type 1 diabetes and the weight-related phenotypes suggest that the role in these reproductive and metabolic phenotypes may likewise trace back to an exocrine pancreatic origin. Significant colocalization analysis (column „Colocalization P-value’, Table 6) calculated using the Simple Sum (1, 72) implemented in LocusFocus (59) between the meconium ileus CF GWAS statistics (1) and the summary statistics from the PheWAS associated traits supports that the same genetic variation contributes to the traits.

**Table 6.**
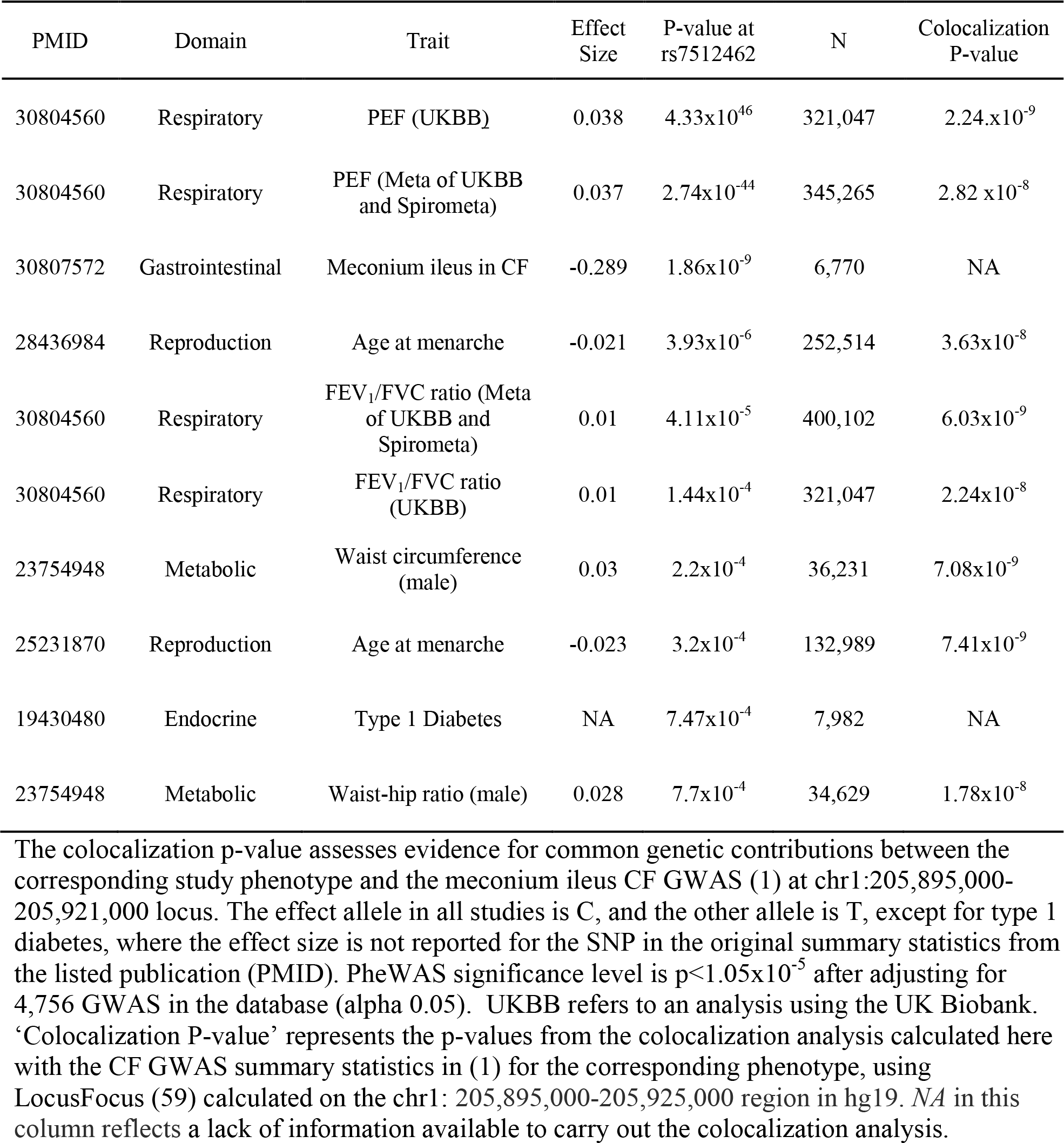
The 10 phenotypes with smallest rs7512462 association p-value obtained from the GWAS ATLAS (41).

| PMID | Domain | Trait | Effect Size | P-value at rs7512462 | N | Colocalization P-value |
| --- | --- | --- | --- | --- | --- | --- |
| 30804560 | Respiratory | PEF (UKBB) | 0.038 | $4.33 \times 10^{-46}$ | 321,047 | $2.24 \times 10^{-9}$ |
| 30804560 | Respiratory | PEF (Meta of UKBB and Spirometa) | 0.037 | $2.74 \times 10^{-44}$ | 345,265 | $2.82 \times 10^{-8}$ |
| 30807572 | Gastrointestinal | Meconium ileus in CF | -0.289 | $1.86 \times 10^{-9}$ | 6,770 | NA |
| 28436984 | Reproduction | Age at menarche | -0.021 | $3.93 \times 10^{-6}$ | 252,514 | $3.63 \times 10^{-8}$ |
| 30804560 | Respiratory | FEV<sub>1</sub>/FVC ratio (Meta of UKBB and Spirometa) | 0.01 | $4.11 \times 10^{-5}$ | 400,102 | $6.03 \times 10^{-9}$ |
| 30804560 | Respiratory | FEV<sub>1</sub>/FVC ratio (UKBB) | 0.01 | $1.44 \times 10^{-4}$ | 321,047 | $2.24 \times 10^{-8}$ |
| 23754948 | Metabolic | Waist circumference (male) | 0.03 | $2.2 \times 10^{-4}$ | 36,231 | $7.08 \times 10^{-9}$ |
| 25231870 | Reproduction | Age at menarche | -0.023 | $3.2 \times 10^{-4}$ | 132,989 | $7.41 \times 10^{-9}$ |
| 19430480 | Endocrine | Type 1 Diabetes | NA | $7.47 \times 10^{-4}$ | 7,982 | NA |
| 23754948 | Metabolic | Waist-hip ratio (male) | 0.028 | $7.7 \times 10^{-4}$ | 34,629 | $1.78 \times 10^{-8}$ |

The colocalization p-value assesses evidence for common genetic contributions between the corresponding study phenotype and the meconium ileus CF GWAS (1) at chr1:205,895,000-205,921,000 locus. The effect allele in all studies is C, and the other allele is T, except for type 1 diabetes, where the effect size is not reported for the SNP in the original summary statistics from the listed publication (PMID). PheWAS significance level is $p < 1.05 \times 10^{-5}$ after adjusting for 4,756 GWAS in the database (alpha 0.05). UKBB refers to an analysis using the UK Biobank. ‘Colocalization P-value’ represents the p-values from the colocalization analysis calculated here with the CF GWAS summary statistics in (1) for the corresponding phenotype, using LocusFocus (59) calculated on the chr1: 205,895,000-205,925,000 region in hg19. NA in this column reflects a lack of information available to carry out the colocalization analysis.

Airflow limitation is a key diagnostic feature of COPD (73). In the UK Biobank we analyzed n=22,071 of 263,461 participants with moderate to severe airflow limitation defined by the Global Initiative for Obstructive Lung Disease (GOLD) criteria (56). Although rs7512462 is a modifier of PEF and FEV_1_/FVC (p=2.74 x10^-44^ and p=4.11×10^-5^, respectively) from ((33); Table 6), the evidence is not as conclusive for FEV_1_pp and COPD case control status in UK Biobank (p=0.56, 0.0891, respectively; Fig. 5).

**Fig. 5.**
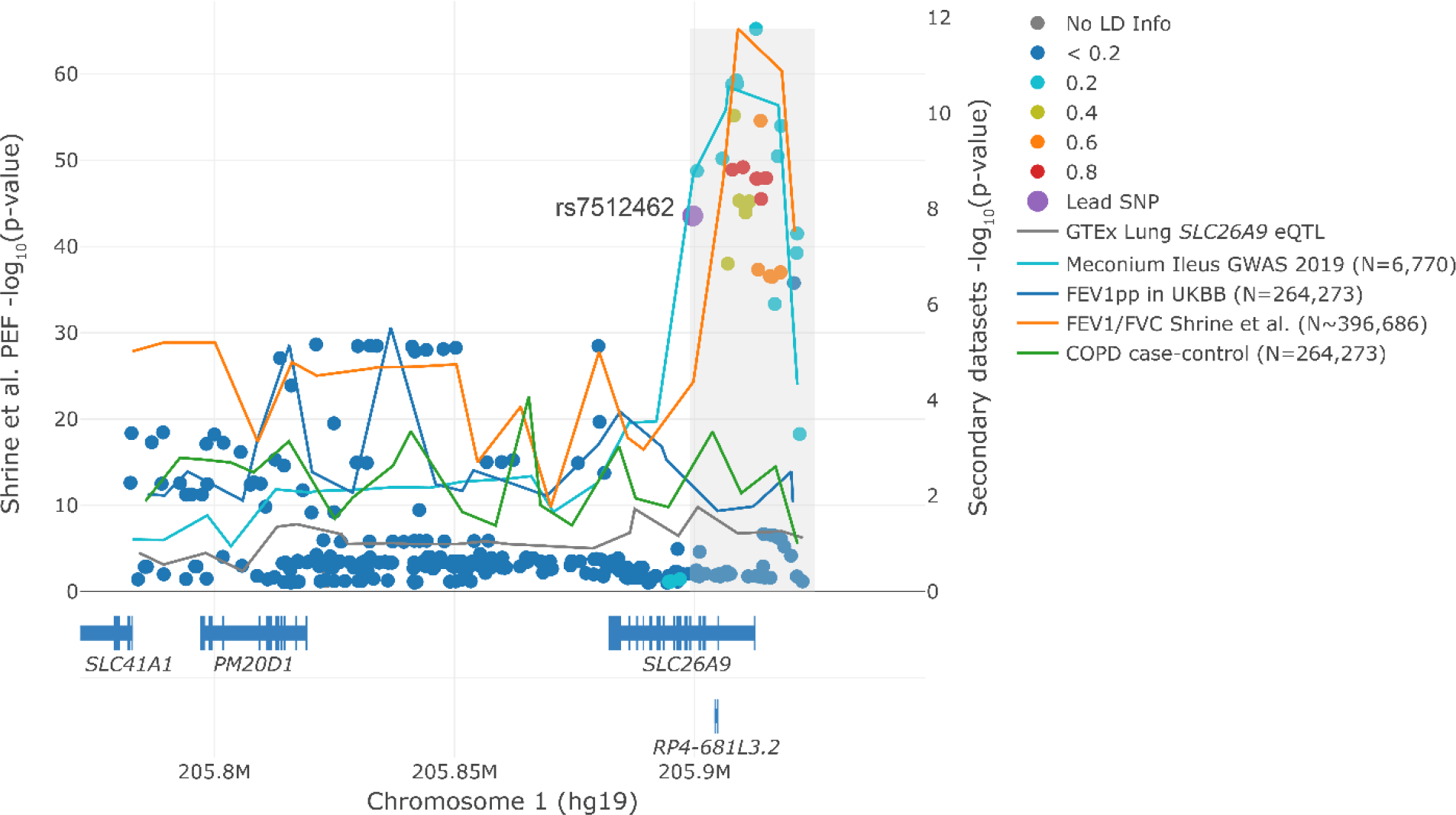
Lung function measures colocalize with CF meconium ileus GWAS at the *SLC26A9* locus. Visualization of colocalization using LocusFocus (59). Peak expiratory flow (PEF) association (n=345,265) from (33) is shown as circles with corresponding left y-axis against lines that trace (1) GTEx v7 adult lung eQTLs for *SLC26A9*; (2) meconium ileus GWAS (1); (3) FEV_1pp_ in UKBB participants (N=263,461; 22,071 of which have COPD as per GOLD level 2-4 definitions); (4) FEV1/FVC ratio from (33) (highest effective sample size is 396,686, a function of sample size (N=400,102) times imputation quality); and (5) Chronic obstructive pulmonary disease (COPD) case-control (22,071 cases and 241,390 controls). The choice of y-axis scales and primary dataset is for visualization purposes only and does not influence colocalization test result conclusions. There is a known gap on the human reference GRCh37 at chr1:205,922,708- 206,072,707, which corresponds to the white gap in the figure as no variants exist. Lines traverse the lowest p-values per window as a visual guide (window size=6.67Kbp) with corresponding right y-axis. A total of 85 SNPs in this region were used to test for colocalization using the Simple Sum method (1, 72). The Simple Sum colocalization test is implemented in the grey region, which was selected to match the observed peak at chr1:205,899,000-205,925,000. The color of the circles reflects the pairwise linkage disequilibrium (LD) with the rs7512462 lead SNP (purple) using the European subset of the 1000 Genomes Project, with varying strength of LD depicted with a different colour as denoted in the legend.

## Discussion

The availability of CFTR modulators is altering care for many individuals with CF, although variation in responses are apparent, partially due to individual genetic backgrounds. One such genetic factor is *SLC26A9,* which contributes to early onset CF comorbidities in the pancreas and intestine (1, 4, 22, 60). Unlike for these early-onset phenotypes where the rs71512462 SNP is a SLC26A9 eQTL in the pancreas and SLC26A9 and CFTR appear to contribute to these phenotypes independently (1, 7, 31), in the lungs the mechanism that is being marked by rs7512462 is not immediately obvious from the genetic data. Meanwhile, some have also questioned whether rs7512462 is even associated with lung function (43).

To address the connection between rs7512462 and SLC26A9, we integrate evidence from population and functional studies. The rs7512462 SNP was shown to associate with lung function or lung response to CFTR-modulators in individuals that carry *CFTR* variants that result in apical membrane localization for CFTR (31) and this is further supported here. Although rs7512462 does not show eQTL evidence based on lung tissue expression, SLC26A9 is present in HBEs (28, 74, 75) and Fig. S3 and Larsen *et al* (28) show that SLC26A9 is a major contributor to constitutive anion secretion across HBEs. In (31) Figure 3B prior to CFTR correction in Phe508del/Phe508del HBE monolayers, rs7512462 shows evidence of association with residual forskolin stimulated current (*P =* 0.09, -0.17 µA/cm^2^ ΔI_eq_-forskolin per C protective allele) in Ussing chamber studies. A relation between rs7512462 genotype and transepithelial current in uncorrected Phe508del CFTR was also shown by others in HNE cultures with Phe508del CFTR genotypes (30).

Experimental evidence for a relationship between the rs7512462 genotype and CFTR function in airway cell models and here in CF participant responses was also observed by rescuing the traffic-defective Phe508del with corrector modulators (30, 31). Published in vitro and cell studies have demonstrated a CFTR-SLC26A9 interaction in lung cells and together with recent work by ((28, 74, 75) and references within), evidence is emerging that SLC26A9 may also stabilize CFTR. Therefore, we interpret that the contribution of SLC26A9 that rs7512462 is marking for lung phenotypes may be from both anion channel activity and enhancement of apical membrane bound CFTR.

To address the uncertainty in evidence for *rs7512462* association with lung function (43), here we extend several genetic association studies. We investigate the role of *rs7512462* genotype in CF patients with different types of CFTR mutations and in treatment response to CFTR modulators both in patient populations and in airway models. We align the findings and previously published work with gene expression patterns including understanding of cell type specific *SLC26A9* and *CFTR* co-expression, and now also consider the role of rs751242 in the phenome and, in particular, in lung function measurements in non-CF populations. Significant human genetic evidence that supports a role for *SLC26A9* in CF disease severity and CFTR modulator response is accumulating. Here we provide the most comprehensive investigation of the role of rs7512462 as a marker of *SLC26A9* activity in CF and non-CF populations.

The relationship between the *SLC26A9* rs7512462 marker and lung function in three types of *CFTR* genotypes was revealing. Individuals with the CF-causing G551D gating variant show association with rs7512462. Gating-deficient CFTR protein exhibits epithelial apical cell membrane localization with reduced opening probability, resulting in reduced epithelial chloride and bicarbonate secretion characteristic of CF (76). In contrast, individuals homozygous for Phe508del, comprising the majority of CF cases, show more modest evidence of association with rs7512462. Phe508del-CFTR is rapidly degraded intracellularly with minimal surface membrane localization. Meanwhile, for the first time, we demonstrated that individuals with MF variants do show rs7512462 association but with absence of CFTR protein cannot respond to CFTR modulator therapy. The studies of MF alleles support that at least some aspect of lung function contribution by SLC26A9 can be independent of CFTR. These investigations align with several previous studies highlighting the potential for SLC26A9 to provide (alternative) chloride transport that functions independently of CFTR (25, 26).

CF GWAS of early-onset CF pancreatic and intestinal phenotypes were CFTR genotype *independent* (1-3, 7); that is, those homozygous for Phe508del still demonstrated genome-wide significance with benefit from the C allele of rs7512462, a marker of *SLC26A9* gene expression in CF phenotypes that show pre-natal onset (for example, meconium ileus). This may reflect that SLC26A9 and CFTR may act independently at this stage of development and is consistent with the observation that *Slc26a9* mRNA was high in the murine pancreas in the embryonic stages of development when *Cftr* was low (31). Given the small sample size, the rs7512462 association with lung function in individuals with two MF alleles requires further investigation and independent replication. However, a consistent beneficial effect of the CC rs7512462 genotype across several of our independent studies and disparate outcomes provides support that modulation of SLC26A9 can provide alternative chloride transport and could be a therapeutic target to improve lung function in individuals with any *CFTR* genotype. The modulation of alternative channels, transporters and pumps to compensate for dysfunctional CFTR (19, 77), and in particular SLC26A9 (9, 20, 31), would provide a mutation-agnostic approach and address the current unmet need of the CF individuals with MF alleles.

Given other published reports assessing the rs7512462 relationship with lung function in individuals with G551D variants (31, 32, 43), we used meta-analysis to summarize the current state of evidence. Together, the weight of evidence supports a relationship between the *SLC26A9* marker, rs7512462, and lung function in individuals with a G551D variant where there is cell-surface localized CFTR protein, but reduced activity. In an expanded CGMS cohort, we also replicated the enhancing effect of the rs7512462 C allele upon rescue of gating variants with IVA, suggesting that the pre-treatment effect may be reflective of constitutive activity and/or attributable to SLC26A9-aided residual CFTR function. Consistent with either is the suggestive association we observed between rs7512462 and lung function prior to treatment with LUM/IVA in individuals from the CGMS homozygous for Phe508del, with a reduced effect size possibly reflective of reduced cell membrane localization and interaction.

LUM/IVA was the first approved modulator for those homozygous for Phe508del, on the basis of reduced pulmonary exacerbations and lung function response. However, the average improvement in lung function in clinical trials (14) and observational studies (34) has been modest. To investigate whether the improvements in lung function from LUM/IVA are modified by the *rs7512462* genotype, we also genotyped the DNA from participants of the US observational LUM/IVA study, PROSPECT (https://clinicaltrials.gov/ct2/show/NCT02477319), and demonstrated that although modest in its average improvement in lung function, those with the CC rs7512462 genotype did exhibit the greatest benefit. Although this is the first report of *rs7512462* impact on clinical response, and in a real-world setting of treated patients, this finding is consistent with published studies of CFTR function in primary HBE and HNE cells from individuals homozygous for Phe508del by us (31) and others (30), respectively. Together these findings across the two treated *CFTR* genotype groups replicate and expand previous reports (25, 26, 30, 31) that rs7512462 correlates with improved CFTR function.

We did evaluate CFTR function in nasal brushes from 36 individuals homozygous for Phe508del in Ussing chamber studies following 24h exposure to vehicle (DMSO), VX-770 with corrector VX-809 (corresponding to IVA/LUM) and a combination of VX-770, VX-809 and an experimental amplifier (68). We observed a similar trend to that reported in (30) with VX-809, although it did not reach statistical significance. When CFTR function was, however, augmented with the amplifier, the difference in CFTR function pre- and post-treatment was greatest in the cells with the rs7512462 CC genotype. These functional studies further support the hypothesis that SLC26A9 will likely benefit any therapeutic situation with increased apical surface localized CFTR protein, such as for the latest highly effective modulator treatment, ETI.

Although the population studies provide important insights into the relationship between CFTR and SLC26A9 marked by rs7512462 *in vivo*, functional studies in cellular and animal models will be necessary to understand the functional relationship between the two channels/transporters (78) and how they may be working together. The expression studies presented here provide guidance on the use of cellular models for SLC26A9 and further highlight potential limitations of cultured HNE for the unique considerations of studying SLC26A9 and CFTR that are distinct from the ones for studies of CFTR alone. Compared to naïve bronchial cells, we observed greater variation and lower expression in naïve nasal cells. Furthermore, the duration of culturing time of either HNE or HBE cells resulted in reduced *SLC26A9* expression. Presently, the cultured HBE model appears superior to HNEs for investigations of SLC26A9, although further investigation of culturing conditions and of cell differentiation should to be considered.

Interestingly, the most significant *SLC26A9* locus SNPs from the CF GWAS (1, 7) also associate with lung function measurements in several large international studies: PEF and FEV_1_/FVC ratio in participants from the UK Biobank aged 40-69 (9, 33); PEF and FEV_1_/FVC ratio in the Spirometa consortium (33); and for the FEV_1_/FVC ratio in 8-year-olds from the UK10K consortium (79). These results align with our findings that, after correction of CF-causal CFTR variants with modulators, *SLC26A9* locus SNPs are associated with improved lung function and CFTR function.

It is noteworthy that decreased spirometry is diagnostic for other obstructive lung diseases such as COPD (80). Several studies have reported similar pathobiology cascades between CF and COPD due to dysfunctional CFTR and environmental risk factors for COPD (81-85). For example, CFTR chloride channels show reduced capacity as a result of tobacco smoke and may result in the mucus obstruction characteristic of COPD (84, 86) that is akin to that seen in CF. If SLC26A9 augments chloride transport, SLC26A9 agonists could also be an effective therapeutic strategy for COPD. In support of this premise is the association evidence provided here demonstrating rs7512462 is a modifier of lung function in (33), with significant colocalization evidence between the CF and their UK Biobank + Spirometa Consortium summary statistics at the *SLC26A9* locus, reflective of a common underlying genetic contribution.

## Conclusions

A role for SLC26A9 in the CF precision medicine landscape is an exciting prospect. SLC26A9 shows desirable characteristics (9) as an alternative therapeutic target for CF, including the urgent need for options for CF individuals with MF alleles. The association between rs7512462 genotype and response to existing pharmaceuticals indicate the potential to refine personalized combinations of modulators, and there is also support that SLC26A9 agonists may yield benefit to any existing pharmacological or gene correction paradigm in CF, independent of *CFTR* genotype. Further research should also address the potential for SLC26A9 agonists to improve measures of lung function in non-CF populations.

## Supporting information

Supplemental File for Genetic evidence supports the development of SLC26A9 targeting therapies including method, result, Supp figure and Supp Tables

## Data Availability

The datasets generated and/or analyzed in this paper are publicly available. Data from the CGMS analyzed for the lung function pre- and post-modulator treatment are available from the Canadian CF registry at https://www.cysticfibrosis.ca/our-programs/cf-registry/requesting-canadian-cf-registry-data; the functional data and RNA-seq data from CGMS is available from the CFIT program at https://lab.research.sickkids.ca/cfit/cystic-fibrosis-patients-families-researchers/, and the paired cultured and fresh naive HNE and HBE is available at GEO (GSE172232). The US PROSPECT data were obtained through application to the US CFF at https://www.cff.org/Research/Researcher-Resources/ and the study is registered on https://clinicaltrials.gov/ct2/show/NCT02477319. The single-cell RNA-sequencing data are downloaded from the Human Lung Cell Atlas at http://hlca.ds.czbiohug.org. The summary statistics for the pheWAS study is available at https://atlas.ctglab.nl/PheWAS and the meconium ileus association results that have been used for colocalization analysis can be downloaded at https://lab.research.sickkids.ca/strug/publications-software/. The data used for the COPD analysis are available through application to the UK Biobank.

## List of abbreviations

CF: cystic fibrosis
CFTR: cystic fibrosis transmembrane conductance regulator
IVA: Ivacaftor
LUM: lumacaftor
TZE: tezacaftor
ETI: elexacaftor and tezacaftor combined with ivacaftor
US: United States
GWAS: genome-wide association study
SLC26A9: Solute Carrier Family 26 member 9
GTEx: Genotype Tissue Expression project
eQTL: expression quantitative locus
HBE: human bronchial epithelia
COPD: chronic obstructive pulmonary disease
CGMS: Canadian CF Gene Modifier Study
HNE: human nasal epithelia
CFF: Cystic Fibrosis Foundation
CCFRD: Canadian CF Patient Data Registry
FEV_1_: forced expiratory volume in one second
MF: minimal function
QC: quality control;FEV_1pp_:forced expiratory volume in one second percent predicted
TPM: transcripts per million
PheWAS: Phenome-wide Association Study
PEF: peak expiratory flow
FVC: the forced vital capacity
FEV_1_/FVC ratio: the ratio of forced expiratory volume in one second to the forced vital capacity
CCFRD: Canadian CF Patient Data Registry
Saknorm: the Survival adjusted average CF-specific Kulich FEV_1_ percentiles that is normalized
QC: quality control
CFIT: CF Canada Sick Kids Program in CF Individualized Therapy
P2: culture at Passage 2
P3: culture at Passage 3
LR: low risk
HR: high risk
PCs: principal components
TMM: trimmed mean of M values
PEER: *probabilistic estimation of expression residual*
RIN: RNA integrity number
ERS/ATS: European Respiratory Society/American Thoracic Society.

## Declarations

### Ethics declarations

The Canadian Gene Modifier Study (CGMS) was approved by the Research Ethics Board of the Hospital for Sick Children (# 0020020214 from 2012-2019 and #1000065760 from 2019- present) and all participating sub-sites. Written informed consent was obtained from all participants or parents/guardians/substitute decision makers prior to inclusion in the study. The CGMS is approved by the Research Ethics Board of the Hospital for Sick Children for the usage of public and external data. The US PROSPECT study provides data from a clinical trial registered at clinicaltrial.gov, identifier NCT02477319, and we obtained these data through application to the US CFF at https://www.cff.org/Research/Researcher-Resources/.

### Consents for publication

Not applicable.

### Data Availability

The datasets generated and/or analyzed in this paper are publicly available. Data from the CGMS analyzed for the lung function pre- and post-modulator treatment are available from the Canadian CF registry at https://www.cysticfibrosis.ca/our-programs/cf-registry/requesting-canadian-cf-registry-data; the functional data and RNA-seq data from CGMS is available from the CFIT program at https://lab.research.sickkids.ca/cfit/cystic-fibrosis-patients-families-researchers/, and the paired cultured and fresh naïve HNE and HBE is available at GEO (GSE172232). The US PROSPECT data were obtained through application to the US CFF at https://www.cff.org/Research/Researcher-Resources/ and the study is registered on https://clinicaltrials.gov/ct2/show/NCT02477319. The single-cell RNA-sequencing data are downloaded from the Human Lung Cell Atlas at http://hlca.ds.czbiohug.org. The summary statistics for the pheWAS study is available at https://atlas.ctglab.nl/PheWAS and the meconium ileus association results that have been used for colocalization analysis can be downloaded at https://lab.research.sickkids.ca/strug/publications-software/. The data used for the COPD analysis are available through application to the UK Biobank.

### Code availability

All code and analyses steps implemented to process the UK Biobank data are available at https://github.com/strug-hub/UKBB_spirometry_on_COPD. The software used for colocalization plotting and statistical testing is available at https://locusfocus.research.sickkids.ca/. The other analyses did not require any custom codes and are described in the Methods Section.

### Competing interests

L.B. participated in a Vertex Virtual Advisory Board and she is a member of the CF Annual Faculty, sponsored by Vertex Pharmaceuticals. D.M-C. received an honorarium for teaching module development for Vertex Pharmaceuticals. N. M. is doing contract research trials for Vertex Phaemaceuticals and Abbvie. A.L.S has received speaking fees for educational programs sponsored by Vertex Pharmaceuticals. B.S.Q. has received speaker fees from Vertex Pharmaceuticals and has served as site PI for several Vertex-sponsored clinical trials .W.M.L is a study investigator for Vertex Pharmaceuticals. E.T., T.G. and F.R. act as a consultant for Vertex Pharmaceuticals. M.S. participated in Vertex clinical trials and received payment for education modules. J.G., G.H., C.W., C. Bartlett, N.P., S.M., F.L., K.K., J.A., A.H., M.S., M.E., G.C-M., D.A., S.B., C.Bjornson, M.C., J.R., A.P., M.P., R.v.W., Y.B., D.H., M.J.S., J.B., P.W., L.S., E.B., T.M., J.M.R and L.J.S. have no conflicts of interest.

### Funding

Funding was provided by Cystic Fibrosis Foundation STRUG17PO, MORAES1610; Canadian Institutes of Health Research (FRN 167282), Cystic Fibrosis Canada (2626) and the CFIT Program funded by the SickKids Foundation and CF Canada; Natural Sciences and Engineering Research Council of Canada (RGPIN-2015- 03742, 250053-2013). This work was also funded by the Government of Canada through Genome Canada (OGI-148) and supported by a grant from the Government of Ontario. The funders of the study play no role in study design, data collection and analysis, decision to publish or preparation of the manuscript.

## Author contributions

L.J.S conceptualized the study and the design. C. Bartlett, F.L., K.K., J.A., A.H., M. Shaw, M.E., G.C-M., D.A., S.B., C. Bjornson, M.C., J.R., A.P., M.P., R.v.W., Y.B., L.B., D.M-C., D.H., M.J.S., N.M., J.B., E.T., A.L.S., B.S.Q., P.W., W.M.L., M. Solomon, E.B., T.J.M., T.G., F.R. enrolled the participants and collected the biospecimens. J.G., G.H., C.W., N.P., S.M., L.J.S. carried out the statistical and bioinformatic analyses, created the visualizations and constructed the tables and Figures. L.S., T.G, J.M.R and L.J.S provided important insights to interpret the results and clarify the technical details for the methods, J.G., G.H., C.W., N.P., J.M.R., L.J.S. wrote the paper and all authors edited the paper. All authors have read and approved the final manuscript.

## Acknowledgments

We thank the US CF Foundation for the use of Part B of the PROSPECT observational study. We thank the Cystic Fibrosis Canada–SickKids Program for Individualized Therapy for access to the RNA-sequencing of the nasal epithelia. We thank the patients, care providers and clinic research assistants, collaborators, and principal investigators involved in CF Centers throughout Canada for their contributions to the CF Canada Patient Registry and Canadian Gene Modifier Study. Some of the research was conducted using the UK Biobank Resource under application Number 40946. We thank Shaf Keshavjee for providing the bronchial cells for paired analysis of gene expression and Hong Ouyang for culturing the epithelial cells. We thank Dr. John Miller for providing the amplifier for the study. The authors wish to acknowledge the staff supporting the High Performance Computing cluster and research helpdesk department and The Centre for Applied Genomics at the Hospital for Sick Children, Toronto.

## Information about Supplementary Material

File name: Supplementary File_clean File format: docx

Title of the data: Supplementary Information; Supplementary Result; Supplementary Figures and Supplementary Tables.

Description of the Data: This file includes the supplementary materials for the paper „**Genetic evidence supports the development of SLC26A9 targeting therapies for the treatment of lung disease*’***

**Supplementary Information** includes the following subsections:

*Sample Genotyping and Quality Control*; *High molecular weight DNA extraction methods; Library preparation and 10x Genomics sequencing reads processing;* Phasing *imputed* genotype

data using *the* 10XG *multi-sample VCF* file as the reference*; HBE and HNE sampling*;

*Cell Culturing; Ussing chamber studies with primary human nasal epithelial cells*; and

*PheWAS data extraction and Colocalization with CF GWAS Summary Statistics*.

**Supplementary Result** includes:

*Haplotype association with Saknorm in CGMS participants homozygous for Phe508del*

There are three figures in section **Supplementary Figures:**

Fig. S1. Forest plot of association between rs7512462 and lung function, measured as Saknorm

(9) in individuals with at least one G551D variant.

Fig. S2. Forest plot of association between rs7512462 and lung function, measured as Saknorm (9) in individuals homozygous for Phe508del.

Fig. S3. SLC26A9 gene expression in various tissue models to guide functional studies.

There are four tables in section **Supplementary Tables**:

**Supplementary Table 1:** Number of participants removed according to the exclusion criteria for the CFTR modulator study.

**Supplementary Table 2:** Meta-analysis for the association between rs7512462 and lung function prior to treatment with Ivacaftor.

**Supplementary Table 3:** Haplotype association with Saknorm in individuals in the CGMS who are homozygous Phe508del.

**Supplementary Table 4:** Association analysis using a linear mixed model with a random intercept with n=45 individuals on ivacaftor with multiple follow-up measures within [15,400] days.

## Notes

### Author Declarations

The Canadian Gene Modifier Study (CGMS) was approved by the Research Ethics Board of the Hospital for Sick Children (# 0020020214 from 2012-2019 and #1000065760 from 2019-present) and all participating sub-sites. Written informed consent was obtained from all participants or parents/guardians/substitute decision makers prior to inclusion in the study. The CGMS is approved by the Research Ethics Board of the Hospital for Sick Children for the usage of public and external data. The US PROSPECT study provides data from a clinical trial registered at clinicaltrial.gov, identifier NCT02477319, and we obtained these data through application to the US CFF at https://www.cff.org/Research/Researcher-Resources/.

### Summary of Updates

In the main manuscript, we redid the meta-analysis for association between rs7512462 and lung function absent of modulator treatment, added a section for haplotype analysis with lung function absent of modulator treatment where haplotype was defined by Lam et al (2020) , and add a linear mixed model for the ivacaftor treatment response using longitudinal data during day 15 to 400 days after treatment. Figure 1 is added for samples size clarification for each substudy in CGMS; Figure 4 is added for the SLC26A9 and CFTR single-cell expression from the Human Lung Cell Atlas to replace the single-cell expression in previous Figure 2 from Human Protein atlas; Figure 2 (original Figure 1) and Figure 5 (original Figure 4) are updated. Part of original Figure 2 is combined with Supplementary Figure 2 to new Figure S3. Table 2 are split to two tables (Table 2 and 3) and a new Table 5 is added for the statistical test for Co-expression between SLC26A9 and CFTR using the Human Lung Atlas data by Zero-Inflated Negative Binomial Model and Spearman's correlation. Supplemental File is also updated.

