## Supplemental File for Genetic evidence supports the development of SLC26A9 targeting therapies including method, result, Supp figure and Supp Tables for "Genetic evidence supports the development of SLC26A9 targeting therapies for the treatment of lung disease"

Program in Genetics and Genome Biology

SickKids Research Institute

Room 12.9705, PGCRL

686 Bay Street

Toronto, ON

M5G 0A4

### Supplementary Information

*Sample Genotyping and Quality Control*

DNA from the CGMS participants (n=2736) were genotyped on four Illumina genome-wide platforms: the 610Quad, 660W, Omni5 (see (1) for details) and Omni2.5 BeadChip. The PROSPECT samples (n=168) were genotyped on the Illumina Omni2.5 BeadChip at The Centre for Applied Genomics, The Hospital for Sick Children. Genotype calling was performed using Genome Studio V2011.1. Quality control (QC) of genotypes were done separately by genotyping platform as in (1) and imputed to a hybrid reference panel consisting of the 1000 Genomes reference and 101 Canadian CGMS participants sequenced at 30X coverage by Complete Genomics (2).

*High molecular weight DNA extraction methods*

High molecular weight (HMW) DNA was extracted from fresh or frozen blood aliquots using the MagAttract HMW DNA Kit (Qiagen, Cat# 67563) as per supplier recommendations. Only samples indicating that bulk DNA was larger than 50kb (>80% by visual inspection of agarose gel) were submitted for sequencing. The full description of the extraction method is provided in (S.M., A. Chen, J.G., F.L., B. Thiruv, W.W. L. Sung, G. Kaur, J. Whitney, Z. Wang, R. V. Patel, K.K., A.H., N.P., J.A., C.W. , G.C-M, D.A., S.B., C.B., M.C., J.R., A.P., M.P., R.v.W., L.B. D.M-C., D.H., M.J.S., N.M., J.B., E.T., A.L.S, B.S.Q, P.W., W.M.L., M.S., E.B. L.S., F.R. L.J.S, High Quality Phasing Using Linked-Read Whole Genome Sequencing of Patient Cohorts Informs Genetic Understanding of Complex Disease, In preparation).

*Library preparation and 10x Genomics sequencing reads processing*

Approximately 1μg of genomic DNA was submitted to The Centre for Applied Genomics at the Hospital for Sick Children for genomic library preparation and whole genome sequencing. After DNA samples were quantified and sample purity was checked, DNA was run on the Genomic Tape on Tapestation (Agilent, Cat# 5067-5365 and 5067-5366) to check DNA fragment size. 10 ng of DNA was used as input material for library preparation using the 10XG Library Kit (PN 120258 and 120257) following the manufacturer's recommended protocol.

Then validated libraries were paired-end sequenced on an Illumina HiSeq X platform following Illumina's recommended protocol to generate paired-end reads of 150-bases in length. The detailed description of library preparation is provided in (S.M., A. Chen, J.G., F.L., B. Thiruv, W.W. L. Sung, G. Kaur, J. Whitney, Z. Wang, R. V. Patel, K.K., A.H., N.P., J.A., C.W. , G.C-M, D.A., S.B., C.B., M.C., J.R., A.P., M.P., R.v.W., L.B. D.M-C., D.H., M.J.S., N.M., J.B., E.T., A.L.S, B.S.Q, P.W., W.M.L., M.S., E.B. L.S., F.R. L.J.S, High Quality Phasing Using Linked-Read Whole Genome Sequencing of Patient Cohorts Informs Genetic Understanding of Complex Disease, In preparation).

Long Ranger 2.2.2 and GRCh38 reference version 2.1.0 were used to process 10XG reads. Base calling was performed using the mkfastq command. VCF files were generated using the wgs command to call and phase variants; GATK 4.0.0.0 was used internally by Long Ranger to call variants (https://support.10xgenomics.com/genome-exome/software/pipelines/latest/what-is-long-ranger). Each 10XG VCF was subset in the region of chr1:205903051-205953456 (GRCh38) and all variants without "PASS" in the FILTER column are set to missing. Bcftools v1.10.2 (3) was used to merge and create a multi-sample VCF for n=4,77 CGMS participants.

*Phasing imputed genotype data using the 10XG multi-sample VCF file as the reference*

SHAPEIT version 4.2.0 (4) was used to completely phase the multi-sample VCF from the 10XG data so that it could be used as a reference panel for imputation in the region chr1:205903051-205953456 (GRCh38). Then SHAPEIT 4.2.0 (4) was used again to phase the multi-sample VCF file from the imputed genotype data of the CGMS (1) using the completely phased VCF from 10XG as the reference panel. LiftoverVCF from picard tools (v2.18.0) is used to lift over the imputed VCF from GRCh38 to GRCh37.

*HBE and HNE sampling*

The HBE samples were collected by bronchoscopy, using a bronchoscopic cytology brush to brush the bronchial airway lumen proximal to the anastomosis and HNE samples were collected by nasal brushing from the inferior turbinate using a 3-mm diameter sterile cytology brush (MP Corporation, Camarillo, CA).

*Cell Culturing*

Cell culturing was carried out in the same manner as described previously (5) for HNE samples from 9 CFIT participants used to investigate expression differences with culturing time (14 versus 28 days); for the 16 paired cultured HNE and HBE samples used to investigate the difference in *SLC26A9* expression across these model systems; and for 46 nasal brushes from individuals homozygous for Phe508del enrolled in the CGMS that were studied in Ussing chamber. Briefly, nasal epithelial cells were isolated and expanded to passage 1 from nasal brushes in the expansion media PneumaCult™ Ex (STEMCELL Technologies) containing 5 µM Rho Kinase inhibitor Y27632 (Selleck Chemicals) and an antibiotic cocktail (penicillin 100 units/mL, streptomycin 100 µg/mL, amphotericin 0.25 µg/mL, tobramycin 80 µg/mL, vancomycin 16 µg/mL, metronidazole 32 µg/mL, meropenem 8 µg/mL, septra (trimethoprim/sulfamethoxazole) 16/80 µg/mL, colistimethate 6 µg/mL). A subset of earlier enrolled samples for Ussing chamber study were cultured using an alternative media using an antibiotic cocktail including penicillin 100 units/mL, streptomycin 100 µg/mL, and amphotericin 0.25 µg/mL. We refer to this alternative media as non-standard media in the analysis.

These expanded cells from homozygous Phe508del samples were seeded to collagen-coated Transwell inserts (6.5 mm diameter, 0.4 µm pore size, Corning) at a seeding density of $1x10^5$ cells per well at passage 2 (P2). Upon confluency, the basolateral media was changed to differentiation media PneumaCult™ ALI (STEMCELL Technologies) containing penicillin 100 units/mL and streptomycin 100 µg/mL. The media was refreshed daily for 7 days then alternate days and any fluid collecting in the apical side was carefully aspirated until the cells reached approximately air-liquid interface (ALI) day 14 for Ussing studies.

Expanded cells for RNA sequencing were further expanded in PneumaCult™ Ex plus (STEMCELL Technologies) media containing the antibiotic cocktail (penicillin 100 units/mL, streptomycin 100 µg/mL, amphotericin 0.25 µg/mL, tobramycin 80 µg/mL, vancomycin 16 µg/mL, metronidazole 32 µg/mL, meropenem 8 µg/mL, septra (trimethoprim/sulfamethoxazole) 16/80 µg/mL, colistimethate 6 µg/mL). Passage 3 cells were seeded in collagen-coated Transwell inserts (6.5 mm diameter, 0.4 µm pore size, Corning) at a seeding density of $1x10^5$ cells per well. The cells were maintained in Pneumacult™ Ex plus media until the Transwells were fully confluent. Upon confluency, the media was changed to differentiation media PneumaCult™ ALI (STEMCELL Technologies). These cells were maintained in an air liquid interface by changing the basolateral media daily for a period of one week, following which the media was changed on alternate days for a period of 14 to 28 days. The 9 HNE samples to study the expression difference at different culture times were sequenced at two time points (14-16 days and 26-30 days) and the paired HNE and HBE samples were well differentiated by 3 weeks before undergoing sequencing.

*Ussing chamber studies with primary human nasal epithelial cells*

Cell monolayers from primary human nasal epithelial (HNE) brushes were mounted in non-perfused P2300 Ussing chambers containing Krebs Bicarbonate buffer (126 mM NaCl, 24 mM $NaHCO_3$, 2.13 mM $K_2HPO_4$, 0.38 mM $KH_2PO_4$, 1 mM $MgSO_4$, 1 mM $CaCl_2$ and 10 mM glucose). The buffer solution was maintained at pH 7.4 and $37^oC$ and continuously gassed with a 5% $CO_2$ / 95% $O_2$ mixture. Transepithelial voltage was recorded using a VCCMC6 amplifier (Physiologic Instruments, San Diego CA) in open-circuit mode and the baseline resistance was measured, following brief 1 μA current pulses every 30 seconds (6) to obtain calculated equivalent short-circuit currents ($I_{eq}$), which was calculated using Ohm's law. Passage 2 confluent cultures were treated with either 0.1% DMSO or CFTR modulators: acute application of Vx770 with 3μM VX-809 (Selleckchem Cedarlane, Canada) or 3μM VX809+1μM of an experimental amplifier (Proteostasis Boston, USA; 42) added to the basolateral ALI media for 24 to 48h (6) prior to Ussing experiments. CFTR function was assessed following inhibition of the epithelial $Na^+$ channel with amiloride (30μM, Spectrum Chemical, Gardena, CA) and following cAMP activation with forskolin (10μM, Sigma-Aldrich, US) in the above treated monolayers. Forskolin-stimulated currents mediated by CFTR were measured as change in current after application of forskolin ($\Delta I_{eq}$-forskolin; $\mu A/cm^2$). The genotype data from the CGMS enabled stratification of CFTR function by rs7512462 genotype to determine whether increased function correlated with genotype upon exposure to the drugs.

*PheWAS data extraction and Colocalization with CF GWAS Summary Statistics*

We extracted all studies with significant p-values at rs7512462 from the GWAS ATLAS at https://atlas.ctglab.nl/PheWAS. By querying the SNP rs7512462 in the 'search SNPs or Gene' box, all the studies that pass the Bonferroni correction are plotted and details about the studies are presented in the lower part of the webpage, which can be downloaded as a csv file from the website. We report the 10 studies with smallest rs7512462 p-value in the paper. The colocalization analysis with the CF GWAS summary statistics (1) was carried out using LocusFocus (v1.4.8) (7).

### Supplementary Result

*Haplotype association with Saknorm in CGMS participants homozygous for Phe508del*

Constructing haplotypes in the CGMS in individuals homozygous for Phe508del as in (60) does not provide evidence for significant low risk (LR) or high risk (HR) haplotypes contributing to Saknorm variation (Supplementary Table 3), although restricting to the subset in the CGMS of individuals with either LR/LR, LR/HR or HR/HR (n=286) does demonstrate significant association evidence (effect size=0.14; p=0.026). Since in this analysis the LR and HR haplotypes are completely tagged by the C and T alleles of rs7512462, the haplotype analysis in this subset of n=286 is equivalent to the association of Saknorm with rs7512462 genotype. Comparing the significant SNP (rs7512462) analysis with the results of the haplotype analysis, we note that although the LR and HR haplotypes are completely tagged, respectively, by the C and T alleles, the reverse is not true. Specifically, the C allele appears on another haplotype, while the T allele appears on five other haplotypes reported in (8). As a result, the HR haplotype analysis uses a comparison haplotype group that includes some haplotypes with the rs7512462 T risk allele and this appears to attenuate the results. The haplotype analysis of this locus akin to that conducted in (8) does not provide greater power than using the single SNP rs7512462 to mark the locus, so all subsequent analyses focus on the rs7512462 genotype.

### Supplementary Figures

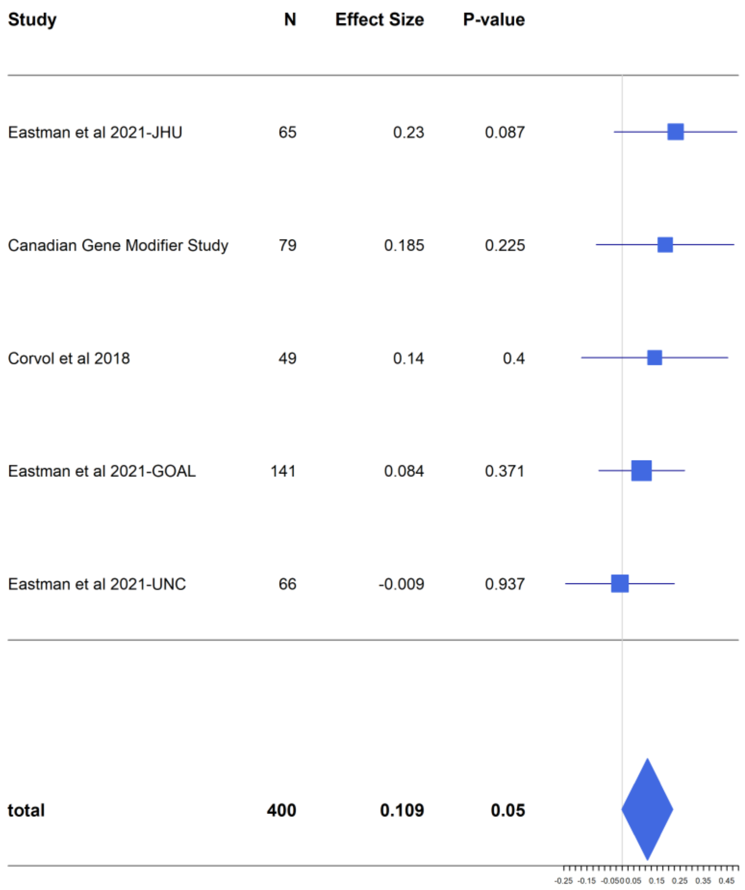


**Fig. S1.** Forest plot of association between rs7512462 and lung function, measured as Saknorm (9) in samples with at least one G551D variant. Saknorm is calculated using $FEV_1$ measured prior to modulator treatment, if applicable. The Canadian Gene Modifier Study (CGMS) association is meta-combined with results from other published studies (10, 11). The CGMS sample here includes 54 individuals included previously in (12) and 25 individuals newly recruited into the CGMS and not previously included in a publication. The inverse variance weighted meta-analysis is reported here.

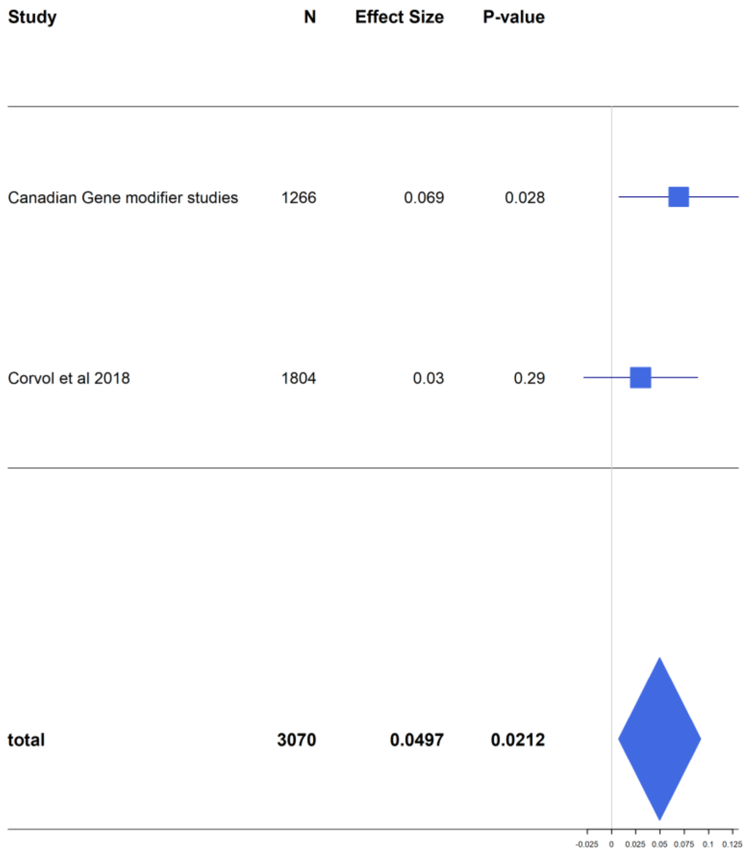


**Fig S2**. Forest plot of association between rs7512462 and lung function , measured as Saknorm (9) in individuals homozygous for Phe508del. Saknorm is calculated using $FEV_1$ measurements prior to modulator treatment, if applicable. The Canadian Gene Modifier Study (CGMS) association is meta-combined with results from the French Gene Modifier Study (10). The CGMS sample here includes 1,013 individuals previously reported in (12) and 253 individuals newly recruited into the CGMS and not previously included in a publication. The result from inverse variance weighted meta-analysis is reported here.

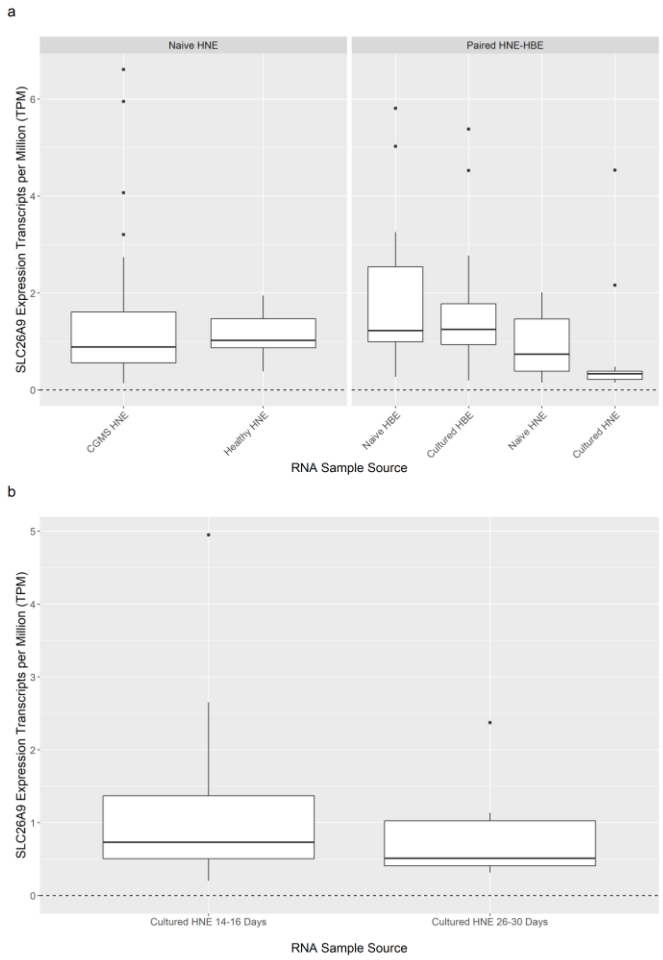


**Fig. S3.** *SLC26A9* gene expression in various tissue models to guide functional studies. (a) *SLC26A9* expression is low in the bulk RNA-Seq of CGMS CF participants and healthy controls in naïve HNE (left), with diminishing expression with culturing of both HNE and HBE (right). (b) *SLC26A9* gene expression in cultured naïve nasal cells from the same individuals (n=9) in CGMS at two different time points. Both sets of cultures were P3.

### Supplementary Tables

**Supplementary Table 1:** Number of participants removed according to the exclusion criteria for the CFTR modulator study. Sample exclusion is in the order of CFTR mutation eligibility for drug, $FEV_{1pp}$ baseline measurement, post-treatment $FEV_{1pp}$ measurement, non-commercial modulator dosage and genotype quality control as shown in the last four columns in the table. All participants included had CFTR genotype approved for the corresponding modulator (at least one gating variant for IVA and homozygous Phe508del for LUM/IVA), and were on the commercial dose, although some of the CGMS participants may have originally gained access to the CFTR modulators through participation in the clinical trial or through compassionate use.

| Studies | CFTR genotype not eligible for drug | $FEV_{1pp}$ baseline missing or measured >3 month of treatment initiation | $FEV_{1pp}$ baseline outside [30,96] | Missing post-treatment $FEV_{1pp}$ measure | Non-commercial modulator dosage | Not genotyped | Genotyping missing rate >10% | Sex mismatch | Non-European ancestry |
|---|---|---|---|---|---|---|---|---|---|
| PROSPECT | 0 | 0 | 58 | 9 | 0 | 0 | 1 | 2 | 7 |
| IVA in CGMS | 15 | 15 | 17 | 1 | 0 | 2 | 0 | 0 | 0 |
| LUM/IVA in CGMS | 2 | 48 | 43 | 67 | 5 | 5 | 0 | 1 | 0 |

**Supplementary Table 2**: Meta-analysis for the association between rs7512462 and lung function prior to treatment with Ivacaftor. Lung function is measured as Saknorm which is a continuous $FEV_1$- based CF-specific percentile that is normalized and accounts for age, sex and height, and is adjusted for survival time (9). Results from inverse variance weighted and sample size weighted meta-analysis are reported.

| | | | | | Inverse-variance weighted meta-analysis | | Sample size weighted meta-analysis | |
|---|---|---|---|---|---|---|---|---|
| CFTR group | Effect size | P-value | N | Study | Effect size | P-value | Effect size | P-value |
| G551D/other | 0.230 | 0.087 | 65 | Eastman et al 2021-JHU | 0.109 | 0.050 | 0.119 | 0.036 |
| | 0.185 | 0.225 | 79 | CGMS | | | | |
| | 0.140 | 0.400 | 49 | Corvol et al 2018 | | | | |
| | 0.084 | 0.371 | 141 | Eastman et al 2021-GOAL | | | | |
| | -0.009 | 0.937 | 66 | Eastman et al 2021-UNC | | | | |
| Phe508del/Phe50del | 0.030 | 0.290 | 1804 | Corvol et al 2018 | 0.050 | 0.021 | 0.046 | 0.037 |
| | 0.069 | 0.028 | 1266 | CGMS | | | | |
| Gating/other | 0.130 | 0.270 | 93 | Corvol et al 2018 | 0.123 | 0.176 | 0.122 | 0.183 |
| | 0.114 | 0.417 | 89 | all CGMS | | | | |

**Supplementary Table 3**. Haplotype association with Saknorm in individuals in the CGMS who are homozygous Phe508del. The haplotypes are constructed in the region chr1: 205899595-205921859 (hg19) as defined in (8) from imputed genotypes (1). The 8 haplotypes defined in (8) are analyzed here, excluding one multi-allelic variant rs144469431.  The first row corresponds to the low risk (LR) haplotype defined in (8) while the last row represents the high risk (HR) haplotype defined in (8). The association results from two analyses are presented: the first is from an analysis that uses the same PLINK command as implemented in  (8)  (PLINK v1.07 with options --chap and --each-vs-others) in 1,164 unrelated CGMS participants with Phe508del/Phe508del and the second is from a linear regression with a robust variance estimator to account for the inclusion of related individuals in 1,266 CGMS participants with Phe508del/Phe508del.

| | Haplotype association CGMS Unrelated Phe508del/Phe508del (n=1164) | | | Haplotype association CGMS Related Phe508del/Phe508del (n=1266) | | |
|---|---|---|---|---|---|---|
| Forty SNP haplotype (chr1: 205899595-205921859) | MHF | Effect size | P-value | MHF | Effect Size | P-value |
| GGCAGCGCGCAAGTGCAATAAGTTCCATATTCCAAGCCCC | 0.267 | (-ref-) | 0.333 | 0.251 | NA | 0.205 |
| GGCAGCGCGCAAGTGCAATAAGCTCCAACGCCCGGGCCCT | 0.047 | 0.107 | 0.121 | 0.042 | 0.152 | 0.027 |
| AGAA-CGGTCAAGTACAATAGACATTGACGTCTGGGCCCC | 0.025 | 0.010 | 0.764 | 0.022 | -0.102 | 0.339 |
| AGAA-CGGTCAAGTACAATAGACATTGACGTCTGGGCCCT | 0.081 | -0.033 | 0.719 | 0.081 | -0.037 | 0.612 |
| AGAAGCGGGCAGT-ACACTAGACATTGACGCCCGGCCCCC | 0.016 | -0.042 | 0.745 | 0.036 | -0.094 | 0.317 |
| AGAA-GGGGCAGT-ATGCAAGACTTTGACGCTCGGGCCCC | 0.073 | -0.025 | 0.791 | 0.064 | 0.014 | 0.740 |
| ACCT-GAGTGGGT-ATGCAAGACATTGACGCCCGGCGTTT | 0.038 | -0.041 | 0.714 | 0.025 | -0.019 | 0.989 |
| ACCT-GAGTGGGT-ATGCACGACATTGACGCTCGGCGTTT | 0.218 | -0.026 | 0.645 | 0.212 | -0.039 | 0.309 |

**Supplementary Table 4.** Association analysis using a linear mixed-effect model with a random intercept with n=45 individuals on ivacaftor with multiple follow-up measures within [15,400] days. Rs7512462 is coded recessively and covariates include $FEV_{1pp}$ and age at baseline, an indicator for whether the participant was in the previous published study ((12); Early CGMS cohort), the number of days between baseline measurement and treatment initiation (Days from baseline measure to treatment), and number of days between treatment initiation and each $FEV_{1pp}$ measurement on treatment (Days from treatment to each treatment measure).

| | Effect size | S. E | t value | P-value |
|---|---|---|---|---|
| rs7512462_CC | 12.759 | 5.471 | 2.332 | 0.025 |
| Fev1pp at baseline | -0.005 | 0.082 | -0.058 | 0.954 |
| Age at baseline | -0.062 | 0.109 | -0.574 | 0.569 |
| Early CGMS cohort | 2.951 | 2.896 | -1.019 | 0.315 |
| Days from baseline measure to treatment initiation | -0.132 | 0.066 | -2.002 | 0.052 |
| Days from treatment to each treatment measure | 0.003 | 0.005 | 0.665 | 0.507 |
